# The re-emergence of influenza following the COVID-19 pandemic in Victoria, Australia

**DOI:** 10.1101/2023.04.02.23288053

**Authors:** Catherine GA Pendrey, Janet Strachan, Heidi Peck, Ammar Aziz, Jean Moselen, Rob Moss, Md Rezanur Rahaman, Ian G. Barr, Kanta Subbarao, Sheena G. Sullivan

## Abstract

**Background:** COVID-19 pandemic mitigation measures, including travel restrictions, effectively limited global circulation of influenza viruses. In Australia, travel bans for non-residents and quarantine requirements for returned travellers were eased in November 2021, providing pathways for influenza viruses to be re-introduced.

**Methods:** From 1 November 2021 to 30 April 2022 we conducted an epidemiological study to investigate the re-establishment of influenza in Victoria, Australia. We analyzed case notification data from the Victorian Department of Health to describe case demographics, interviewed the first 200 cases to establish probable routes of virus reintroduction, and examined phylogenetic and antigenic data to understand virus diversity and susceptibility to current vaccines.

**Results:** Overall, 1598 notifications and 1064 positive specimens were analyzed. The majority of cases occurred in the 15-34 year age group. Case interviews revealed a higher incidence of international travel exposure during the first month of case detections and high levels of transmission in university residential colleges associated with the return to campus. Influenza A(H3N2) was the dominant subtype, with a single lineage predominating despite multiple importations.

**Conclusions:** Enhanced testing for respiratory viruses during the COVID-19 pandemic provided a more complete picture of influenza virus transmission compared to previous seasons. Returned international travellers were important drivers of the re-emergence of influenza, as were young adults, a group whose role has previously been under-recognised in the establishment of seasonal influenza epidemics. Targeting interventions, including vaccination, to these groups could reduce influenza transmission in the future.

## Introduction

Although recently overshadowed by the impact of the COVID-19 pandemic, influenza remains an important cause of global morbidity and mortality. Seasonal influenza is estimated to cause 290,000-650,000 respiratory deaths each year, in addition to deaths from cardiovascular and other causes [1].

In temperate climates, influenza epidemiology is characterised by seasonal epidemics during the cooler months, while in tropical regions outbreaks can occur year-round [2]. However, during 2020 and 2021 measures to control the COVID-19 pandemic resulted in unprecedented global suppression of circulating influenza viruses [3, 4]. In the World Health Organization European region there was a 99.8% reduction in the number of influenza specimens testing positive from October 2020 to February 2021, despite high levels of influenza testing, following the introduction of strict COVID-19 pandemic control measures (eg. mask wearing and limitations on gatherings) [5]. Similar patterns were seen in countries around the world [3], as influenza outbreaks were largely confined to parts of Asia and West Africa during this period [6].

In Australia, as part of the COVID-19 pandemic response, the federal government introduced progressive travel restrictions beginning in late February 2020, culminating in border closure to all non-residents on 20 March 2020 and introduction of 14-days mandatory quarantine in supervised facilities (principally, hotels) on 28 March 2020 [7]. Initiation of travel restrictions coincided with a dramatic reduction in influenza in Australia [8]. Only 33 of 60,031 specimens tested in Australia were positive for influenza between April-July 2020 [3], and between April 2020 and October 2021, confirmed cases were largely confined to quarantined travellers [8].

The reduction in global influenza transmission was accompanied by a decrease in genetic diversity (bottlenecks) and marked geographic differentiation. Bottlenecks occurred for A(H1N1), A(H3N2) and B/Victoria viruses [4], and in the most extreme example, B/Yamagata was not detected after April 2020 [9]. Phylogenetic differentiation of influenza viruses was observed to mirror the geographic boundaries of travel restrictions in West Africa [4].

On the 1^st^ November 2021 Australia lifted quarantine requirements for COVID-19-vaccinated travellers entering into some jurisdictions, including Victoria [10]. The reopening of national borders and reinvigoration of international travel provided an opportunity for influenza viruses to recirculate [11]. These unique circumstances provided an ideal setting to study the establishment of seasonal influenza epidemics. We aimed to prospectively characterise the epidemiological and virological dynamics of influenza’s re-emergence in Victoria to enhance understanding of seasonal epidemics in Australia and comparable countries and inform future public health responses.

## Methods

Here, we describe the reintroduction of influenza to Victoria for the 6-month period between 1 November 2021, when quarantine-free international travel resumed, and 30 April 2021, by which time localized transmission in Victoria was clear. Victoria is located in the southern temperate region of Australia, and has a population of 6.5 million, predominantly residing in Melbourne, a city of 4.9 million [12]. Influenza is notifiable in Victoria (and throughout Australia) with all cases of laboratory-confirmed influenza required to be notified to the Victorian Department of Health (DoH) by the diagnosing laboratories under the Public Health and Wellbeing Act 2008 [13]. Under an enhanced surveillance protocol, laboratories were requested to send influenza-positive specimens to the World Health Organization Collaborating Centre for Reference and Research on Influenza (WHO CCRRI) in Melbourne, for further characterisation, and the first 200 confirmed cases were contacted for case interviews, as described below.

### Case notifications

Case notification data for laboratory-confirmed influenza were extracted from the DoH Public Health Event Surveillance System. These included basic demographic information (age, sex, location of residence by Statistical Area Level 1 (SA1)) and influenza sample information (collection date, type and subtype). Residential SA1 was used to approximate the socioeconomic position of cases using 2016 Australian Census data, measured by the index of relative social advantage and disadvantage (IRSAD) [14]. Data were examined for demographic trends. Case notifications were also compared with COVID-19 case notification numbers, using data accessed from publicly available data on the Victorian DoH website [15]. Descriptive analyses were performed using R for Windows 4.1.2.

### Antigenic and genetic analyses

Positive influenza specimens were forwarded to WHO CCRRI for genetic and antigenic characterisation. Virus culture was attempted for all specimens in MDCK-SIAT1 cells (Madin-Darby canine kidney cells transfected with human 2,6-sialyltransferase) [16]. Antigenic similarity to the 2022 southern hemisphere vaccine viruses was assessed by haemagglutination inhibition (HI) assay, as previously described [17]. This assay measured the ability of post infection ferret antisera raised against representative viruses from the 2022 southern hemisphere vaccines - egg- and cell-grown A/Victoria/2570/2019 (H1N1)pdm09-like virus, and cell-grown A/Darwin/6/2021 (H3N2)-like virus to inhibit viral binding of red blood cells [9].

Sequencing was performed on the haemagglutinin (HA) gene from virus isolates or original specimens if an isolate was not available, using Sanger or Illumina iSeq as previously described [18]. Phylogenetic analysis and tree construction was undertaken using the maximum likelihood method and Augur pipeline [19], which uses IQTree [20] for constructing and bootstrapping (-B 1000 -alrt 1000) the phylogenetic tree (model: GTR), then visualised using the R package ggtree [21].

### Case interviews

We identified the first 200 confirmed cases of influenza notified to the DoH after 1 November 2021 for interview. This approach was based on the First Few X protocols, originally used in the 2009 pandemic [22] and later adopted by the WHO [23] to characterize the epidemiological, clinical and virological properties of emerging respiratory pathogens through follow up of early cases and contacts. From 31 March to 10 May 2022 we attempted to contact each confirmed case for interview. The standard DoH influenza interview form was used and amended to include questions on pathways to access influenza testing, in addition to existing items addressing vaccination status, risk factors for severe disease (including medical co-morbidity and pregnancy) and exposure to travel, agricultural settings, and influenza cases. Clusters were identified based on epidemiological linkages (common exposure sites) and compared to phylogenetic data, where available, to identify potential transmission events.

### Ethics

This follow up of cases was conducted under the auspices of the Public Health and Wellbeing Act (Victoria, 2008) [13] and associated Public Health and Wellbeing Regulations (2019) [24]. The collection, use and disclosure of personal information was in accordance with the Privacy and Data Protection Act 2014 (Vic) and the Health Records Act 2001 (Vic). Ethical approval for analyses of these data was provided by the ANU Human Research Ethics Committee (2022/208).

## Results

### Influenza case notifications

The first influenza notification was on 5 December 2021. Numbers remained low over the summer (December-February), with 41 notifications, including only three in February 2022 (Figure 1A). This drop coincided with a SARS-CoV-2 epidemic wave (Figure 1B). Regular influenza notifications progressively increased during March with a sustained upsurge that continued into April 2022. By the end of April, 1598 laboratory-confirmed cases had been notified to DoH, all being diagnosed as Influenza A. Based on information submitted by diagnostic laboratories, subtype was only available for 87 cases (5.4%), of which 85 (97.7%) were A(H3N2) and two (2.3%) were A(H1N1)pdm09. By comparison, there were 1,478,163 COVID-19 cases in Victoria over the same period (Figure 1B).

**Figure 1.**
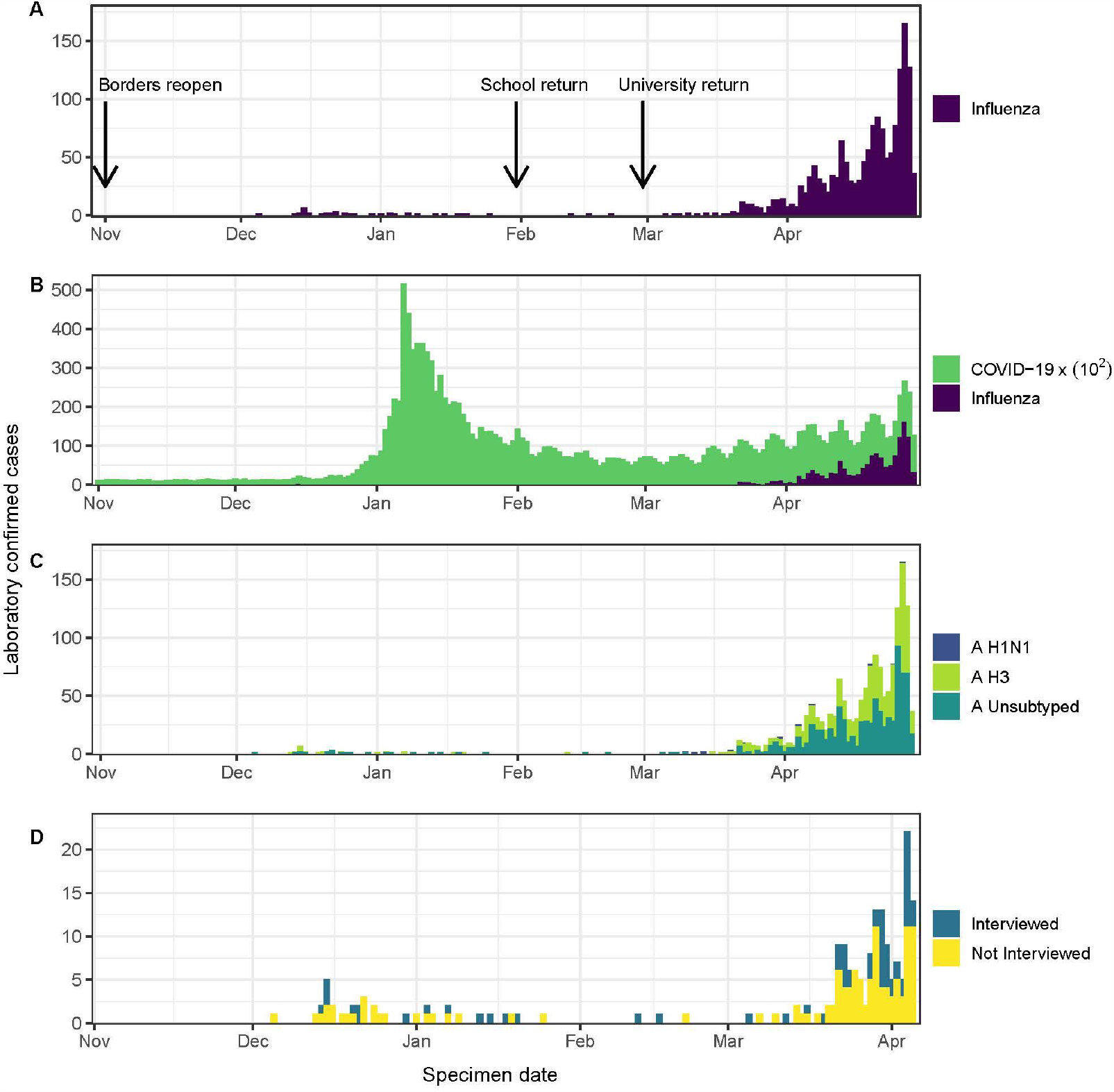
Confirmed influenza cases, Victoria, Australia, 1 November 2021 – 30 April 2022 (A) Influenza cases; (B) Influenza cases by subtype; (C) Influenza and COVID-19 cases and (D) first 200 influenza cases by interview by completion status. Figure 1 notes: Panel A: Source: Victorian Department of Health. Panel B. Source: Victorian Department of Health and WHO Collaborating Centre for Reference and Research on Influenza. Panel C: Source: Victorian Department of Health. Panel D: Daily influenza case notifications for first 200 case notifications 1 November 2021 to 6 April 2022. Source: Victoria Department of Health and study interview data.

There was a slight predominance of female cases (Figure 2). Nearly half of cases were young adults aged 15-24 years (22.6% aged 20-24 years and 20.7% aged 15-19 years; Supplementary Table 1). Individuals in age groups at increased risk of severe influenza comprised 15.4% of cases, 6.0% <5 years, and 9.4% ≥65 years. In March when cases began to increase, the 20-24 year old age group dominated with 41/108 cases (38.0%), followed by 15-19 year olds with 25/108 cases (23.1%). In April the predominance of those aged 15-24 years partially abated, as the relative proportion of cases increased in all other age groups, except individuals aged ≥75 years.

**Figure 2.**
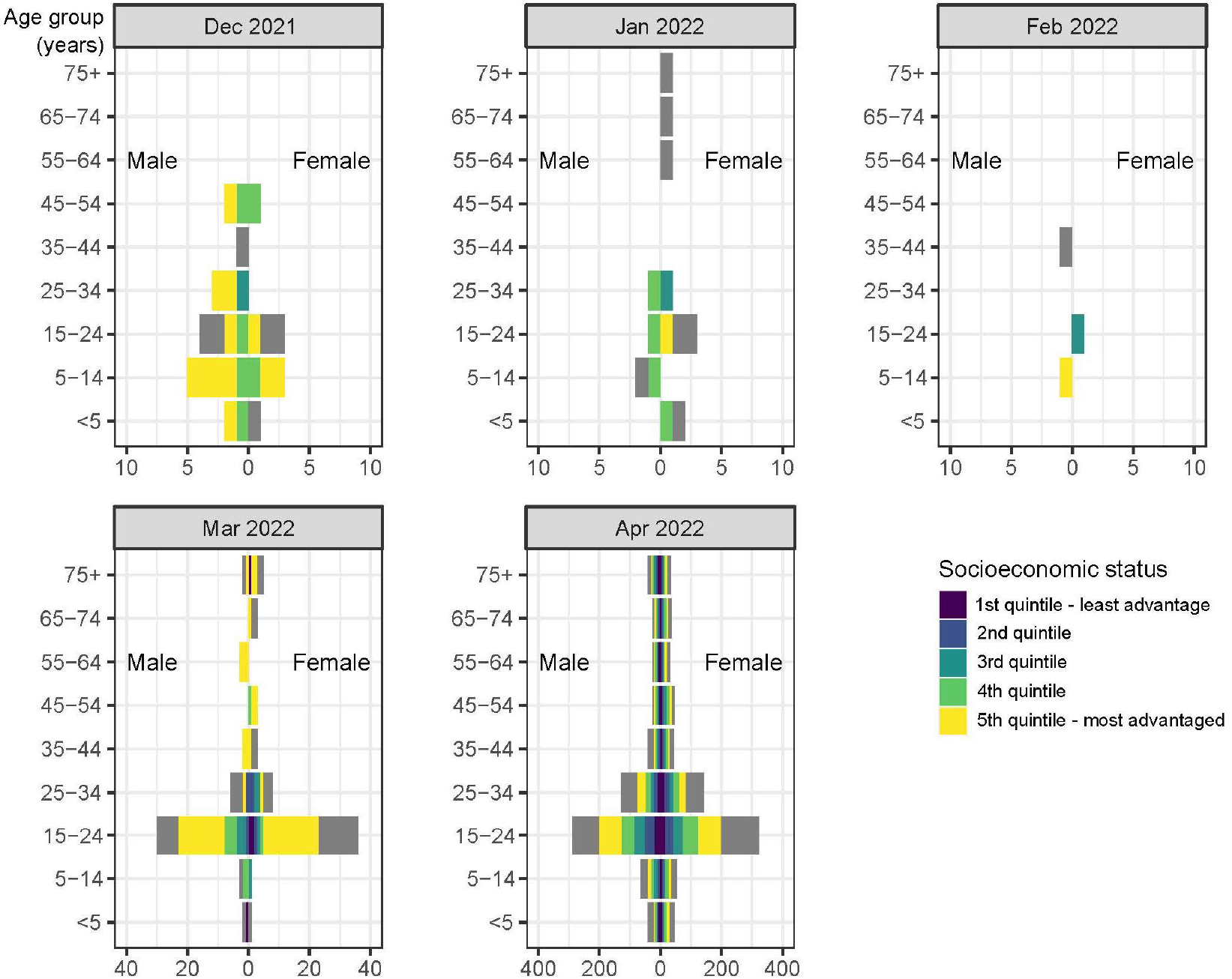
Demographic characteristics of influenza cases by month of infection, Victoria, Australia, 1 November 2021 – 30 April 2022 Figure 2 notes: Socioeconomic status according to the index of relative social advantage and disadvantage (IRSAD), based on location of residence. Source: Victorian Department of Health and Australian Bureau of Statistics.

Of the 1087 cases with available remoteness area classification, 84.4% occurred in major cities, 13.2% in inner regional areas and 2.4% in outer regional areas. Of the 1058 cases with an available socio-economic indicator, 13.2% were in the lowest quintile for Victoria and 33.5% were in the highest. The skew towards relative advantage was more pronounced prior to the increase in cases in April. From November 2021-March 2022 61/98 (62.2%) cases lived in highest quintile areas while only 4/98 (4.1%) lived in lowest quintile areas.

### Antigenic analysis

From 1 November 2021 until 30 April 2022, 1064 positive influenza samples were sent to WHO CCRRI from Victorian diagnostic laboratories. The age and sex distribution of individuals with positive samples was similar to DoH notification data (Supplementary Table 2). All samples were influenza A, with no influenza B viruses identified (Figure 1C). Subtype was available for 714 samples (67.1%), of which 699 (97.9%) were A(H3N2), and just 15 (2.1%) were A(H1N1)pdm09.

Of 1064 samples, HI was performed on 645 (60.6%) virus isolates. Of 633 A(H3N2) isolates, 607 (95.9%) were antigenically similar to A/Darwin/6/2021. All 12 A(H1N1)pdm09 virus isolates demonstrated a similar antigenic profile to the A/Victoria/2570/2019 virus.

### Genomic sequencing

Of all specimens sent to WHO CCRRI, whole genome sequencing results were available for 192 (18.0%). Of 179 A(H3N2) viruses, 178 (99.4%) fell into the same HA genetic clade as the vaccine virus (A/Darwin/6/2021), the 3C.2a1b.2a.2 clade [9]. All A(H1N1) influenza viruses sequenced were within the same HA clade as the vaccine virus (A/Victoria/2570/2019), that is 6B.1A.5a.2 (see Supplementary table 3).

A HA phylogenetic tree was constructed using 175/192 A(H3N2) viruses, with 17 excluded due to incomplete sequences and low depth coverage. The phylogenetic tree revealed a dominant genetic group within the HA 3C.2a1b.2a.2 clade with high levels of homogeneity, indicating a probable point source outbreak (Figure 3)

**Figure 3.**
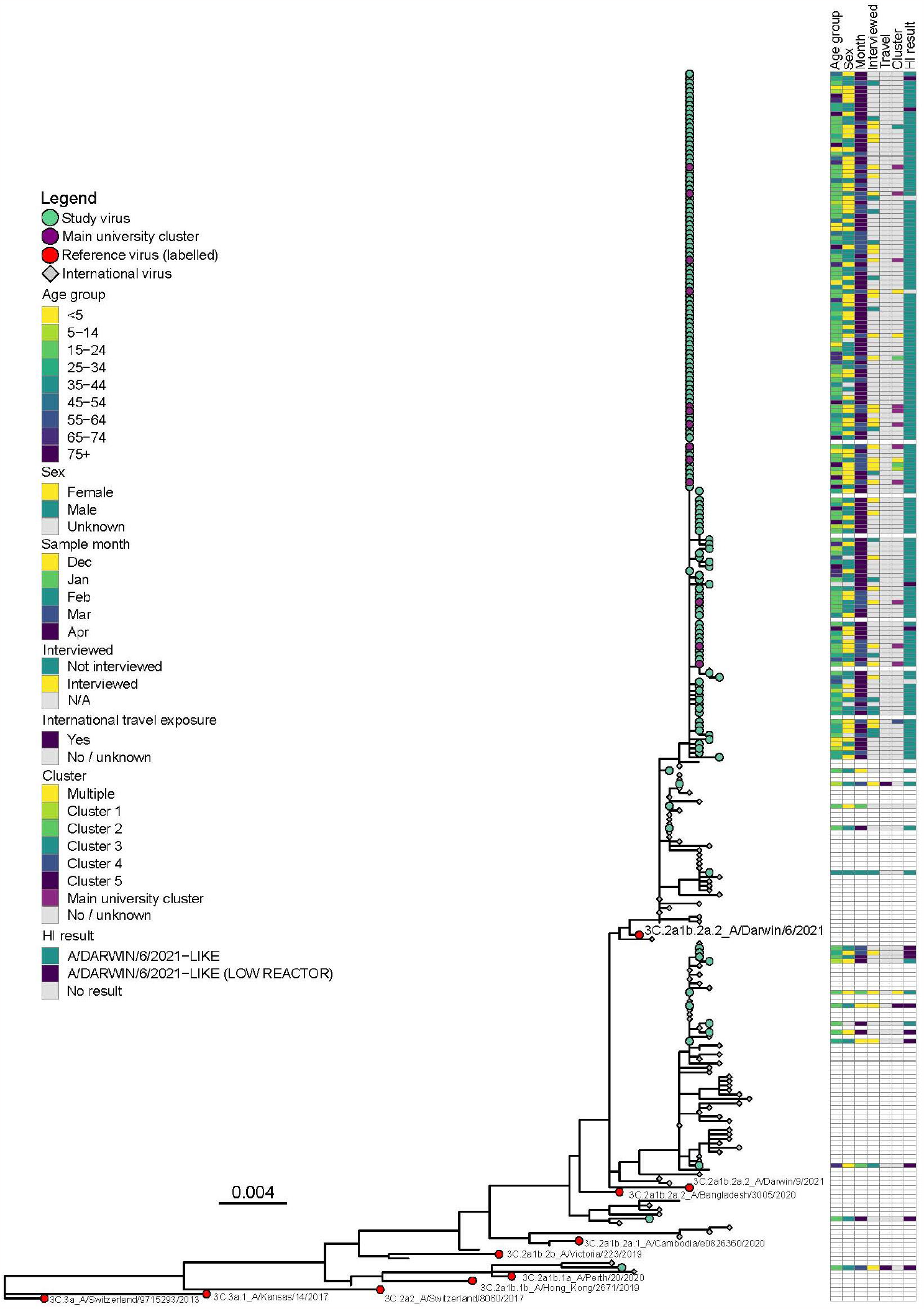
Phylogenetic trees of A(H3N2) influenza viruses, HA gene, Victoria, Australia, 1 Nov 2021 - 30 Apr 2022. Figure 3 notes: GISAID accession numbers and information available for viruses sequenced in this study in Supplementary table 3 and for international and reference virus in Supplementary table

### Case interviews

The first 200 case notifications were reported to DoH between 5 December 2021 and 6 April 2022. Of these 178 (89.0%) had available contact information and 124 (62.0%) were successfully contacted for interview (Figure 1D). There were no significant differences in the age and sex distributions, between those who were and were not interviewed (Supplementary Table 5).

Of those who were interviewed, the median age was 20.9 years (interquartile range 18.8-27.6) (Table 1). There were approximately even numbers of male and female cases with the vast majority (116/124, 93.5%) living in major cities, and the remainder in inner regional areas.

**Table 1.**
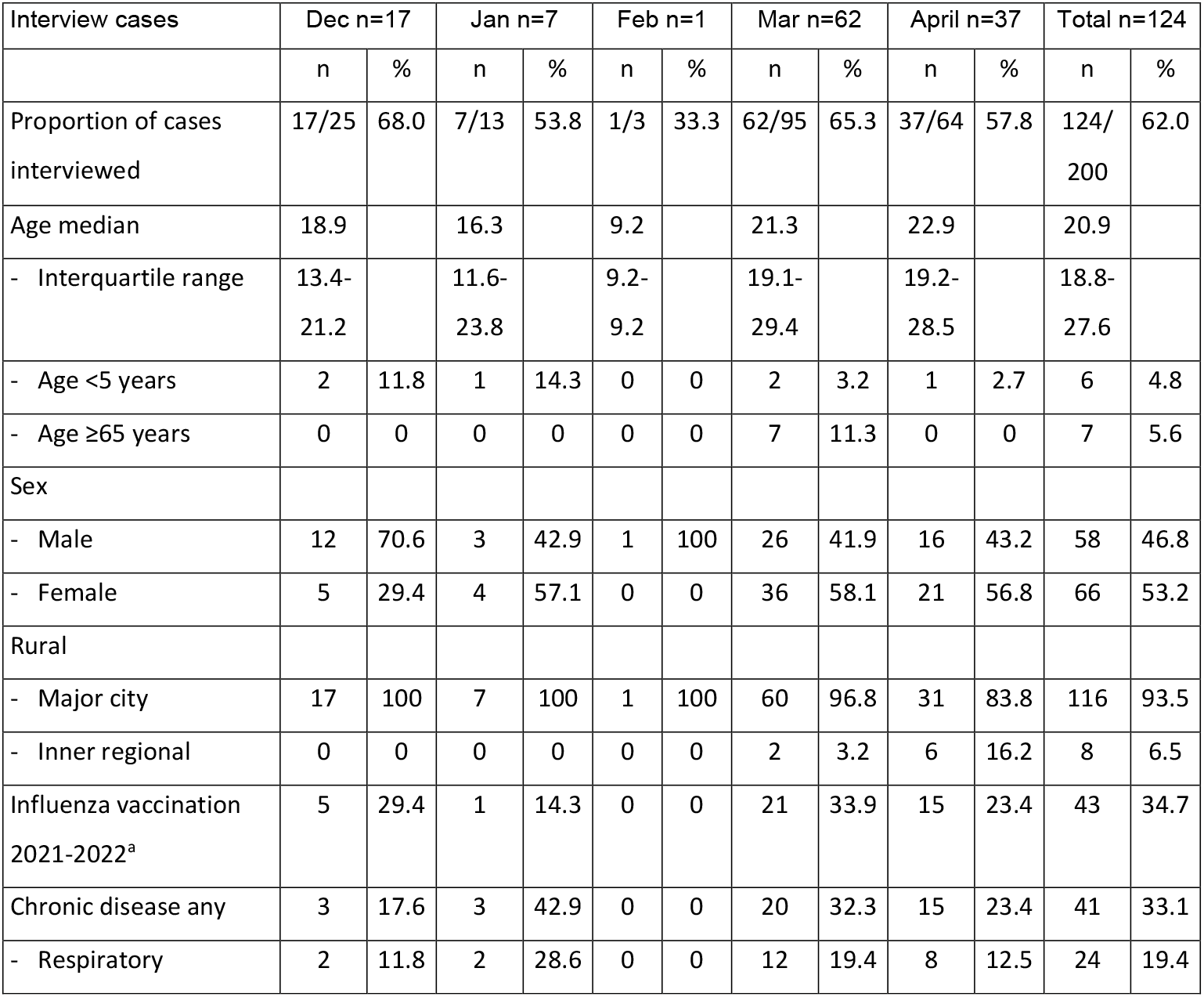
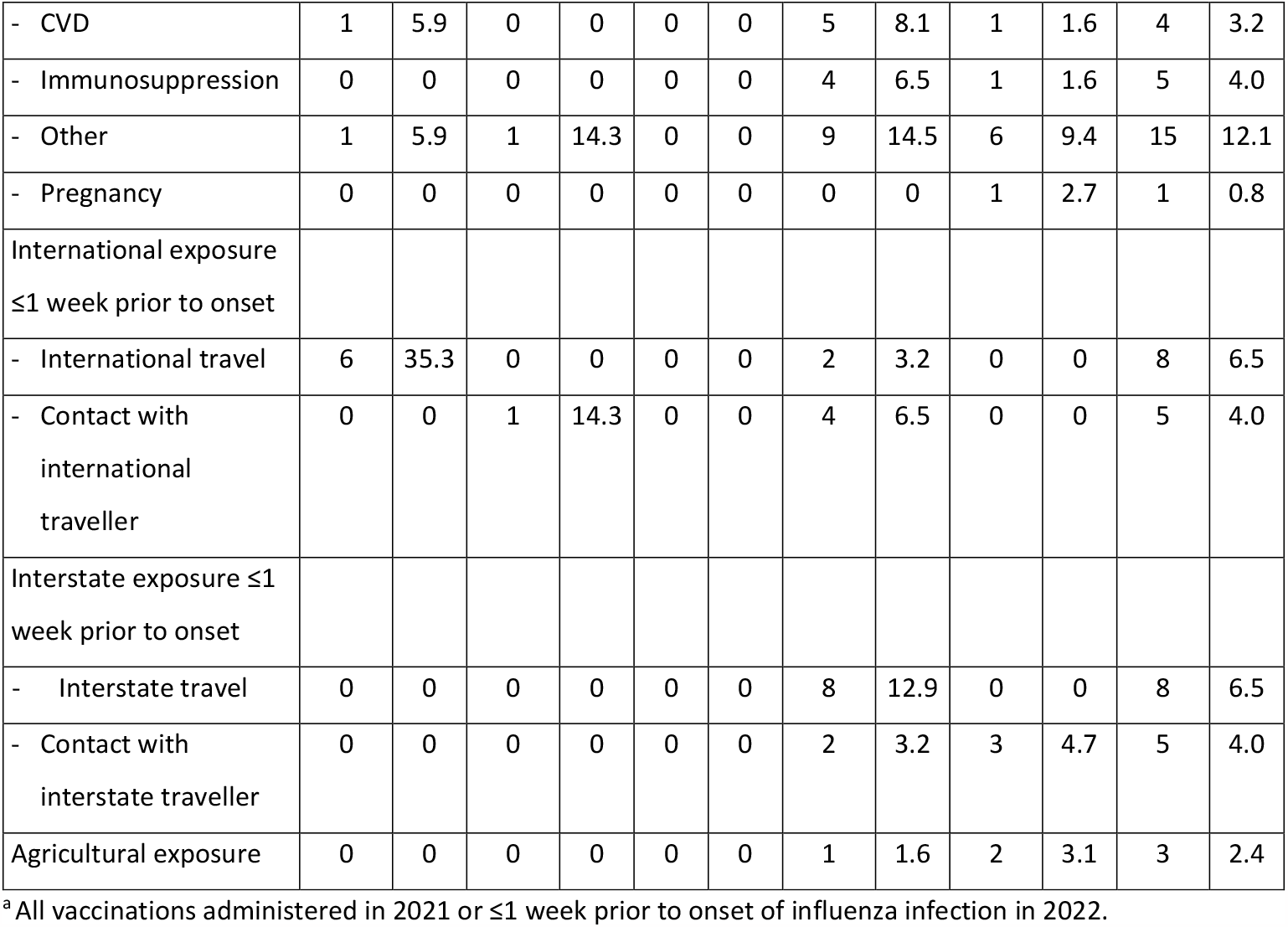
Characteristics of interviewed influenza cases, 1 Nov 2021 - 6 Apr 2022, Victoria

| Interview cases | Dec n=17 |  | Jan n=7 |  | Feb n=1 |  | Mar n=62 |  | April n=37 |  | Total n=124 |  |
| --- | --- | --- | --- | --- | --- | --- | --- | --- | --- | --- | --- | --- |
|  | n | % | n | % | n | % | n | % | n | % | n | % |
| Proportion of cases interviewed | 17/25 | 68.0 | 7/13 | 53.8 | 1/3 | 33.3 | 62/95 | 65.3 | 37/64 | 57.8 | 124/200 | 62.0 |
| Age median | 18.9 |  | 16.3 |  | 9.2 |  | 21.3 |  | 22.9 |  | 20.9 |  |
| - Interquartile range | 13.4-21.2 |  | 11.6-23.8 |  | 9.2-9.2 |  | 19.1-29.4 |  | 19.2-28.5 |  | 18.8-27.6 |  |
| - Age <5 years | 2 | 11.8 | 1 | 14.3 | 0 | 0 | 2 | 3.2 | 1 | 2.7 | 6 | 4.8 |
| - Age ≥65 years | 0 | 0 | 0 | 0 | 0 | 0 | 7 | 11.3 | 0 | 0 | 7 | 5.6 |
| Sex |  |  |  |  |  |  |  |  |  |  |  |  |
| - Male | 12 | 70.6 | 3 | 42.9 | 1 | 100 | 26 | 41.9 | 16 | 43.2 | 58 | 46.8 |
| - Female | 5 | 29.4 | 4 | 57.1 | 0 | 0 | 36 | 58.1 | 21 | 56.8 | 66 | 53.2 |
| Rural |  |  |  |  |  |  |  |  |  |  |  |  |
| - Major city | 17 | 100 | 7 | 100 | 1 | 100 | 60 | 96.8 | 31 | 83.8 | 116 | 93.5 |
| - Inner regional | 0 | 0 | 0 | 0 | 0 | 0 | 2 | 3.2 | 6 | 16.2 | 8 | 6.5 |
| Influenza vaccination 2021-2022<sup>a</sup> | 5 | 29.4 | 1 | 14.3 | 0 | 0 | 21 | 33.9 | 15 | 23.4 | 43 | 34.7 |
| Chronic disease any | 3 | 17.6 | 3 | 42.9 | 0 | 0 | 20 | 32.3 | 15 | 23.4 | 41 | 33.1 |
| - Respiratory | 2 | 11.8 | 2 | 28.6 | 0 | 0 | 12 | 19.4 | 8 | 12.5 | 24 | 19.4 |
| - CVD | 1 | 5.9 | 0 | 0 | 0 | 0 | 5 | 8.1 | 1 | 1.6 | 4 | 3.2 |
| - Immunosuppression | 0 | 0 | 0 | 0 | 0 | 0 | 4 | 6.5 | 1 | 1.6 | 5 | 4.0 |
| - Other | 1 | 5.9 | 1 | 14.3 | 0 | 0 | 9 | 14.5 | 6 | 9.4 | 15 | 12.1 |
| - Pregnancy | 0 | 0 | 0 | 0 | 0 | 0 | 0 | 0 | 1 | 2.7 | 1 | 0.8 |
| International exposure<br>≤1 week prior to onset |  |  |  |  |  |  |  |  |  |  |  |  |
| - International travel | 6 | 35.3 | 0 | 0 | 0 | 0 | 2 | 3.2 | 0 | 0 | 8 | 6.5 |
| - Contact with<br>international<br>traveller | 0 | 0 | 1 | 14.3 | 0 | 0 | 4 | 6.5 | 0 | 0 | 5 | 4.0 |
| Interstate exposure ≤1<br>week prior to onset |  |  |  |  |  |  |  |  |  |  |  |  |
| - Interstate travel | 0 | 0 | 0 | 0 | 0 | 0 | 8 | 12.9 | 0 | 0 | 8 | 6.5 |
| - Contact with<br>interstate traveller | 0 | 0 | 0 | 0 | 0 | 0 | 2 | 3.2 | 3 | 4.7 | 5 | 4.0 |
| Agricultural exposure | 0 | 0 | 0 | 0 | 0 | 0 | 1 | 1.6 | 2 | 3.1 | 3 | 2.4 |

<sup>a</sup> All vaccinations administered in 2021 or ≤1 week prior to onset of influenza infection in 2022.

Cases were interviewed about exposures within one week of symptom onset. Of the 124 interviewed cases, eight (6.5%) reported recent international travel, including six in December 2021. An additional five cases reported close contact with a returned international traveller, one in January and four in March. Uncertainty of the representativeness of these proportions was higher in months with lower numbers of interviewed cases (Supplementary Figure 1). Thirteen cases (10.5%) reported a history of interstate travel or close contact with a returned interstate traveller, all testing positive between 21 March and 6 April 2022. Only three cases reported spending time in an agricultural setting, including one reporting poultry contact.

Among interviewees, 41 reported having received influenza vaccination in 2021 and seven in 2022 (including one individual also vaccinated in 2021). The date of all 2022 vaccinations was reported as either unknown or ≤1 week prior to influenza onset. One third of interviewed cases reported having a chronic medical condition. Respiratory conditions were the most common (19.4%) and included a diagnosis of asthma in all cases.

Regarding health service pathways to accessing influenza testing, primary care clinics were the most common location of first attendances (45.1%) and presentations (54.1%), followed by emergency departments (first attendances: 24.6%; presentations: 45.9%) and COVID-19 testing clinics (first attendances 20.5%; presentations: 21.3%) (Supplementary Table 6). Accordingly, more than 20% of interviewed cases secondarily presented to an emergency department after attending another health service within their episode of illness to seek medical review.

Through the limited contact and exposure history obtained, 56 cases (45.2%) were linked to nine clusters (Supplementary Figure 2). Remaining cases had no identified epidemiological links to other cases. There was one dominant cluster of 30 people associated with a single university campus that coincided with the increase in case detections in March. Within this cluster 12 cases lived in onsite university residential accommodation. During interviews, these cases described widespread transmission of influenza and acute respiratory illness other than COVID-19 on campus and especially within residential colleges. An additional seven cases were associated with other universities. Secondary transmission was reported within households and through social contacts. Other clusters with multiple cases were associated with music festivals, schools, private functions and hospitality venues.

Comparison of epidemiological and phylogenetic data indicated that all thirteen of the cases with sequences identified as being within the main university epidemiological cluster were part of the dominant lineage within the A(H3N2) HA 3C.2a1b.2a2 clade (Figure 3). The two cases with both sequences and an identified history of interstate travel were also within this dominant genetic group. In contrast, neither of the two individuals with identified international travel exposure and sequences were within this genetic group. The A(H3N2) virus from one international traveller from Somalia was the only sequence identified within the HA 3C.2a1b.1a clade.

## Discussion

After an 18-month period of effective local elimination of influenza in Australia, mainly through travel restrictions and enforced quarantining, positive influenza case detections began shortly after the recommencement of quarantine-free international travel in November 2021. Case numbers remained low during the summer holiday period, coinciding with a SARS-CoV-2 (Omicron BA.1) epidemic, and then showed a steady increase of A(H3N2) from late March, which was possibly accelerated in universities. Increased access to multi-pathogen testing during the COVID-19 pandemic has enabled testing among groups not normally captured in surveillance data [25], which has probably provided a more complete picture of influenza spread in the community in 2022 compared with previous years.

Consistent with previous studies, our findings highlighted the important role of travel-related importations in establishing seasonal influenza epidemics [26-28]. The frequency of international arrivals among interviewed cases (6.5%) exceeded the rate observed in the Australian population over the same period, equivalent to 1.1% per month from December 2021–April 2022 [12, 29]. We also identified a higher rate of international travel exposure earlier in the study period, 35.3% in December compared to 6.5% in January-April. Although numbers were small, together with the re-commencement of cases after border opening, this suggests importations led to sustained local transmission. A study from New South Wales (NSW), Australia, during the 2018-19 summer also identified the importance of travel during summer epidemics, with influenza cases seven times more likely to report recent international travel exposure during the first two months of their study, compared to later in the summer [26]. Belderok et al. similarly identified the importance of travel-related importations in a European setting in a 2006-2007 study, with 7.2% of short-term travellers to tropical and subtropical regions contracting influenza, and half of those with symptomatic influenza infection considered contagious on return to the Netherlands [27].

Previous phylogenetic studies have determined that influenza virus circulation in Australia is seeded by multiple importations from the global virus population, followed by rapid local dissemination [28]. In our study, we observed multiple incursions, but overwhelming predominance of a single A(H3N2) 3C.2a1b.2a.2 subgroup. The genetically homogenous grouping could represent a point-source outbreak or multiple introductions of genetically-similar viruses, as has previously been observed in university-based influenza surveillance [30]. Reduced transmission during the summer, competition with the first Omicron epidemic and ongoing adherence with NPIs as well as incomplete sampling, and stochastic transmission variability, may all account for the limited observed onward transmission of some viruses [31].

Our study highlighted the potential importance of young adults in establishing seasonal influenza epidemics. During March, as case numbers started to increase, and universities returned to onsite teaching, almost 40% of cases occurred in adults aged 20-24 years. The influenza literature has traditionally emphasized the role of school-aged children in transmission, rather than young adults [32]. However, notable influenza outbreaks on university campuses since relaxation of pandemic travel restrictions [33, 34], and evidence from the COVID-19 pandemic that community-wide transmission was predicted by increased disease incidence among people aged <25 years [35], underscore the important—and previously underappreciated—role of young adults in transmission. Young adults have high levels of mobility and social interactions [36, 37], but comparatively lower rates of influenza vaccination [38], and lower use of NPIs [39]. Younger individuals, with less cumulative exposure to influenza, may also have been disproportionately vulnerable to influenza infection after two years of no transmission [40].

Universities were identified in interviews as an important setting for transmission. Numerous university outbreaks also occurred in the USA early in the post-pandemic re-emergence of influenza, including a rapidly proliferating A(H3N2) outbreak at the University of Michigan [33, 34]. One modelling study concluded that university campuses are susceptible to rapid transmission and high attack rates during influenza outbreaks. University residential colleges increase risks, as high-density accommodation and shared facilities promote inter-personal contact and constrain the capacity to limit transmission [37]. The return to university, after the recommencement of international travel during the holiday period, likely compounded the risk of outbreaks in Melbourne. From November 2021 there was a dramatic increase in the total number of overseas arrivals to Australia. Over the same period the number of international students arriving in Australia also rebounded [29]. International students have restricted access to subsidised healthcare (including influenza vaccination and testing, but were not excluded from free SARS-CoV-2 testing or vaccination), potentially increasing transmission risks for this group. Some universities offer free seasonal influenza vaccination, but this is not universally available.

The high proportion of young adults and international travellers in our study raises the prospect of targeting interventions to these groups to reduce the overall burden of influenza. Vaccination is the most practical means of mitigating influenza epidemics and is provided free in Australia under the National Immunisation Program (NIP) to high risk groups including children <5 years old, adults ≥65 years old, individuals with specified chronic medical conditions, Aboriginal and Torres Strait Islander individuals and pregnant women [41]. Individuals who are not eligible may access influenza vaccination privately but at their own cost. In June 2022, in response to concern that rapidly-rising influenza case numbers could overwhelm health systems already strained by the pandemic, eligibility for free influenza vaccination was expanded to all age groups in most Australian jurisdictions, including Victoria. In future seasons, maintaining expanded eligibility for subsidized vaccination and promoting vaccination among travellers could reduce the community burden of influenza. A similar approach, in which vaccination is provided free to school-age children, has been employed in the United Kingdom [42], Western Australia [43], and Japan [44]. Marsh et al. also highlighted the importance of promoting vaccination among travellers, including revaccination for those vaccinated earlier in the season, to mitigate importations propagating outbreaks [26]. Specifically promoting vaccination and testing among young adults may also be an effective strategy to reduce overall community transmission. Likewise, maintaining some level of NPIs for acute respiratory illnesses has the potential to reduce influenza-related morbidity and mortality [45].

A key mitigation measure used in Australia during the COVID-19 pandemic was testing to guide isolation [46], with results communicated directly to those being tested, rather than via clinicians as was previously the case. During our study it was apparent that some cases were not aware of their influenza diagnosis, which limits a case’s ability to isolate to protect others. Following this study, the government has worked with pathology providers to communicate influenza and other respiratory virus test results direct to consumers. As influenza is often viewed as more severe than the ‘common cold,’ it is hoped that receiving positive results will increase the likelihood individuals adopt respiratory hygiene measures [39, 47].

The 2022 influenza season in Australia progressed after our study ended and was characterized by an unusually early start, a sharp rise in cases, followed by a rapid decline, and a moderately severe epidemic season overall [48]. The season commenced prior to the annual influenza vaccination campaign in early April, by which time population immunity from the previous year’s vaccine was waning [49]. Influenza case numbers peaked in late May and early June during the Omicron BA.2 epidemic, and proceeded to rapidly decline from late June. Expansion in influenza case numbers was accompanied by an increase in the diversity of influenza viruses in Victoria, including detections of B/Victoria viruses and increased diversity in A(H3N2) viruses [9]. There was some variability across Australia. Several jurisdictions (NSW, Northern Territory and Western Australia) had higher proportions of A(H1N1) early in the season [11]. However, A(H3N2) was ultimately the dominant subtype in all jurisdictions, many in the same dominant genetic subgroup as observed in our study.

### Limitations

It is unlikely that our study captured all international importations of influenza, which would include asymptomatic and mild cases that were not tested for influenza. It is possible that under-ascertainment could have been greater early in the study period, prior to broad recognition that influenza had returned. Genetic sequences were available for 18.0% of positive specimens. More complete sequencing may have helped to provide a richer understanding of transmission dynamics. Incomplete recollection of events limited our ability to identify epidemiological links between cases.

## Conclusions

We have documented the re-establishment of endemic influenza in Victoria, Australia, after a prolonged absence of circulation. Our findings highlight the importance of international travellers and young adults, in promoting the spread of influenza viruses, and the potential value of targeting interventions to these groups. As we have now reconnected globally, efforts to increase influenza vaccination uptake should continue to be prioritized, with increased focus on young adults and international travellers.

## Data Availability

All data produced in the present work are contained in the manuscript.

## Notes

### Competing Interest Statement

SGS reports honoraria from CSL Seqirus and Novavax.
IGB owns shares in a vaccine producing company.
Remaining authors have no conflicts to declare.

### Funding Statement

The WHO Collaborating Centre for Reference and Research on Influenza is supported by the Australian Government Department of Health and Aged Care.

### Author Declarations

The Human Research Ethics Committee of the Australian National University gave ethical approval for this work (data analysis). Case follow up was conducted under the auspices of the Public Health and Wellbeing Act (2008) and associated Public Health and Wellbeing Regulations (2019) of the Victorian Government.

