## Supplementary material for "The re-emergence of influenza following the COVID-19 pandemic in Victoria, Australia"

Supplementary table 1. Demographic characteristics of influenza cases, Victoria, Australia, 1 November 2021 – 30 April 2022.

| | 2021-12 (n=25) | | 2022-01 (n=13) | | 2022-02 (n=3) | | 2022-03 (n=108) | | 2022-04 (n=1449) | | Total (n=1598) | |
|---|---|---|---|---|---|---|---|---|---|---|---|---|
| Sex | n | % | n | % | n | % | n | % | n | % | n | % |
| - Female | 8 | 32.0 | 9 | 69.2 | 1 | 33.3 | 60 | 55.6 | 761 | 52.5 | 839 | 52.5 |
| - Male | 17 | 68.0 | 4 | 30.8 | 2 | 66.7 | 48 | 44.4 | 683 | 47.1 | 754 | 47.2 |
| - Not stated | 0 | 0.0 | 0 | 0.0 | 0 | 0.0 | 0 | 0.0 | 5 | 0.3 | 5 | 0.3 |
| Age group (years) | | | | | | | | | | | | |
| - <5 | 3 | 12.0 | 2 | 15.4 | 0 | 0.0 | 3 | 2.8 | 88 | 6.1 | 96 | 6.0 |
| - 5-10 | 1 | 4.0 | 2 | 15.4 | 1 | 33.3 | 3 | 2.8 | 64 | 4.4 | 71 | 4.4 |
| - 10-14 | 7 | 28.0 | 0 | 0.0 | 0 | 0.0 | 1 | 0.9 | 57 | 3.9 | 65 | 4.1 |
| - 15-19 | 3 | 12.0 | 3 | 23.1 | 1 | 33.3 | 25 | 23.1 | 298 | 20.6 | 330 | 20.7 |
| - 20-24 | 4 | 16.0 | 1 | 7.7 | 0 | 0.0 | 41 | 38.0 | 315 | 21.7 | 361 | 22.6 |
| - 25-29 | 2 | 8.0 | 2 | 15.4 | 0 | 0.0 | 10 | 9.3 | 164 | 11.3 | 178 | 11.1 |
| - 30-34 | 1 | 4.0 | 0 | 0.0 | 0 | 0.0 | 4 | 3.7 | 107 | 7.4 | 112 | 7.0 |
| - 35-44 | 1 | 4.0 | 0 | 0.0 | 1 | 33.3 | 5 | 4.6 | 86 | 5.9 | 93 | 5.8 |
| - 45-54 | 3 | 12.0 | 0 | 0.0 | 0 | 0.0 | 3 | 2.8 | 74 | 5.1 | 80 | 5.0 |
| - 55-64 | 0 | 0.0 | 1 | 7.7 | 0 | 0.0 | 3 | 2.8 | 58 | 4.0 | 62 | 3.9 |
| - 65-74 | 0 | 0.0 | 1 | 7.7 | 0 | 0.0 | 3 | 2.8 | 62 | 4.3 | 66 | 4.1 |
| - ≥75 | 0 | 0.0 | 1 | 7.7 | 0 | 0.0 | 7 | 6.5 | 76 | 5.2 | 84 | 5.3 |
| Rurality | | | | | | | | | | | | |
| - Major City | 17 | 68.0 | 6 | 46.2 | 2 | 66.7 | 77 | 71.3 | 815 | 56.2 | 917 | 57.4 |
| - Inner Regional | 2 | 8.0 | 0 | 0.0 | 0 | 0.0 | 2 | 1.9 | 140 | 9.7 | 144 | 9.0 |
| - Outer Regional | 0 | 0.0 | 0 | 0.0 | 0 | 0.0 | 0 | 0.0 | 26 | 1.8 | 26 | 1.6 |
| - Unknown | 6 | 24.0 | 7 | 53.8 | 1 | 33.3 | 29 | 26.9 | 468 | 32.3 | 511 | 32.0 |
| Socioeconomic status [a] | | | | | | | | | | | | |
| - 1st quintile | 0 | 0.0 | 0 | 0.0 | 0 | 0.0 | 4 | 3.7 | 136 | 9.4 | 140 | 8.8 |
| - 2nd quintile | 0 | 0.0 | 0 | 0.0 | 0 | 0.0 | 5 | 4.6 | 154 | 10.7 | 159 | 10.0 |
| - 3rd quintile | 1 | 4.0 | 1 | 7.7 | 1 | 33.3 | 7 | 6.5 | 167 | 11.6 | 177 | 11.1 |
| - 4th quintile | 6 | 24.0 | 4 | 30.8 | 0 | 0.0 | 8 | 7.4 | 210 | 14.5 | 228 | 14.3 |
| - 5th quintile | 12 | 48.0 | 1 | 7.7 | 1 | 33.3 | 47 | 43.5 | 293 | 20.3 | 354 | 22.2 |
| - Unknown | 6 | 24.0 | 7 | 53.8 | 1 | 33.3 | 37 | 34.3 | 484 | 33.5 | 535 | 33.6 |

[a] Socioeconomic status according to the index of relative social advantage and disadvantage (IRSAD), based on location of residence, 1st quintile represents the least advantaged area and 5th quintile the most advantaged area.
Data source: Victorian Department of Health

Supplementary Table 2. Comparison between demographics of influenza cases reported to the Department of Health and cases with a specimen available for antigenic and genetic analysis at WHO CCRRI

| | WHO CCRRI specimens (n=1064 | | Department of Health cases (n=1598 | | p-value[a] |
|---|---|---|---|---|---|
| Sex | | | | | 0.66 |
| - Female | 419 | 39.4% | 839 | 52.5% | |
| - Male | 361 | 33.9% | 754 | 47.2% | |
| - Unknown | 284 | 26.7% | 5 | 0.3% | |
| Age group | | | | | 0.54 |
| - <5 | 59 | 5.5% | 96 | 6.0% | |
| - 5-14 | 82 | 7.7% | 136 | 8.5% | |
| - 15-24 | 416 | 39.1% | 691 | 43.2% | |
| - 25-34 | 188 | 17.7% | 290 | 18.1% | |
| - 35-44 | 69 | 6.5% | 93 | 5.8% | |
| - 45-54 | 51 | 4.8% | 80 | 5.0% | |
| - 55-64 | 52 | 4.9% | 62 | 3.9% | |
| - 65-74 | 54 | 5.1% | 66 | 4.1% | |
| - ≥75 | 64 | 6.0% | 84 | 5.3% | |
| - Unknown | 29 | 2.7% | 0 | 0.0% | |

[a]p-values calculated with chi-squared tests, excluding unknown values. Based on inclusion of unknown values, p=<0.001 for both age and sex.

Supplementary Table 3. GISAID Accession numbers for influenza viruses sequenced, Victoria, Australia, 1 November 2021 – 30 April 2022

| Isolate_Id | Isolate_Name | Clade | Subtype | Submitting_Lab | Authors | Originating_Lab |
|---|---|---|---|---|---|---|
| EPI_ISL_11499449 | A/Victoria/2/2022 | 3C.2a1b.2a.2 | A/H3N2 | WHO CCRRI | This study | Monash Medical Centre |
| EPI_ISL_12004355 | A/Victoria/24/2022 | 3C.2a1b.2a.2 | A/H3N2 | WHO CCRRI | This study | ST Vincent Hospital |
| EPI_ISL_12004532 | A/Victoria/26/2022 | 3C.2a1b.2a.2 | A/H3N2 | WHO CCRRI | This study | Australian Clinical Laboratories |
| EPI_ISL_12004533 | A/Victoria/27/2022 | 3C.2a1b.2a.2 | A/H3N2 | WHO CCRRI | This study | ST Vincent Hospital |
| EPI_ISL_12004534 | A/Victoria/17/2021 | 3C.2a1b.2a.2 | A/H3N2 | WHO CCRRI | This study | Alfred Hospital |
| EPI_ISL_12206246 | A/Victoria/6/2022 | 3C.2a1b.2a.2 | A/H3N2 | WHO CCRRI | This study | Alfred Hospital |
| EPI_ISL_12206250 | A/Victoria/12/2022 | 3C.2a1b.2a.2 | A/H3N2 | WHO CCRRI | This study | Royal Melbourne Hospital |
| EPI_ISL_12206251 | A/Victoria/10/2022 | 3C.2a1b.2a.2 | A/H3N2 | WHO CCRRI | This study | Alfred Hospital |
| EPI_ISL_12206253 | A/Victoria/5/2022 | 3C.2a1b.2a.2 | A/H3N2 | WHO CCRRI | This study | Alfred Hospital |
| EPI_ISL_12206256 | A/Victoria/16/2022 | 3C.2a1b.2a.2 | A/H3N2 | WHO CCRRI | This study | Royal Melbourne Hospital |
| EPI_ISL_12206257 | A/Victoria/18/2022 | 3C.2a1b.2a.2 | A/H3N2 | WHO CCRRI | This study | Royal Melbourne Hospital |
| EPI_ISL_12206258 | A/Victoria/17/2022 | 3C.2a1b.2a.2 | A/H3N2 | WHO CCRRI | This study | Royal Melbourne Hospital |
| EPI_ISL_12206259 | A/Victoria/11/2022 | 3C.2a1b.2a.2 | A/H3N2 | WHO CCRRI | This study | Alfred Hospital |
| EPI_ISL_12206260 | A/Victoria/15/2022 | 3C.2a1b.2a.2 | A/H3N2 | WHO CCRRI | This study | Monash Medical Centre |
| EPI_ISL_12206264 | A/Victoria/4/2022 | 3C.2a1b.2a.2 | A/H3N2 | WHO CCRRI | This study | Alfred Hospital |
| EPI_ISL_12206274 | A/Victoria/7/2022 | 6B.1A.5a.2 | A/H1N1 | WHO CCRRI | This study | WHO CCRRI |
| EPI_ISL_12206276 | A/Victoria/13/2022 | 6B.1A.5a.2 | A/H1N1 | WHO CCRRI | This study | Monash Medical Centre |
| EPI_ISL_12206280 | A/Victoria/9/2022 | 6B.1A.5a.2 | A/H1N1 | WHO CCRRI | This study | WHO CCRRI |
| EPI_ISL_12579823 | A/Victoria/47/2022 | 3C.2a1b.2a.2 | A/H3N2 | WHO CCRRI | This study | ST Vincent Hospital |
| EPI_ISL_12579824 | A/Victoria/51/2022 | 3C.2a1b.2a.2 | A/H3N2 | WHO CCRRI | This study | Dorevtich Pathology |
| EPI_ISL_12579835 | A/Victoria/52/2022 | 3C.2a1b.2a.2 | A/H3N2 | WHO CCRRI | This study | Dorevtich Pathology |
| EPI_ISL_12579850 | A/Victoria/53/2022 | 3C.2a1b.2a.2 | A/H3N2 | WHO CCRRI | This study | Dorevtich Pathology |
| EPI_ISL_12579866 | A/Victoria/54/2022 | 3C.2a1b.2a.2 | A/H3N2 | WHO CCRRI | This study | Dorevtich Pathology |
| EPI_ISL_12579868 | A/Victoria/56/2022 | 3C.2a1b.2a.2 | A/H3N2 | WHO CCRRI | This study | ST Vincent Hospital |
| EPI_ISL_12579870 | A/Victoria/66/2022 | 6B.1A.5a.2 | A/H1N1 | WHO CCRRI | This study | Royal Melbourne Hospital |
| EPI_ISL_12579871 | A/Victoria/49/2022 | 3C.2a1b.2a.2 | A/H3N2 | WHO CCRRI | This study | Royal Melbourne Hospital |
| EPI_ISL_12579872 | A/Victoria/50/2022 | 3C.2a1b.2a.2 | A/H3N2 | WHO CCRRI | This study | Dorevtich Pathology |
| EPI_ISL_12579875 | A/Victoria/67a/2022 | 3C.2a1b.2a.2 | A/H3N2 | WHO CCRRI | This study | Royal Melbourne Hospital |
| EPI_ISL_12579876 | A/Victoria/42/2022 | 3C.2a1b.2a.2 | A/H3N2 | WHO CCRRI | This study | Alfred Hospital |
| EPI_ISL_12579877 | A/Victoria/62/2022 | 3C.2a1b.2a.2 | A/H3N2 | WHO CCRRI | This study | Monash Medical Centre |
| EPI_ISL_12579878 | A/Victoria/123/2022 | 3C.2a1b.2a.2 | A/H3N2 | WHO CCRRI | This study | Royal Melbourne Hospital |
| EPI_ISL_12579879 | A/Victoria/124/2022 | 3C.2a1b.2a.2 | A/H3N2 | WHO CCRRI | This study | Royal Melbourne Hospital |
| EPI_ISL_12579880 | A/Victoria/67/2022 | 3C.2a1b.2a.2 | A/H3N2 | WHO CCRRI | This study | Royal Melbourne Hospital |
| EPI_ISL_12579881 | A/Victoria/99/2022 | 3C.2a1b.2a.2 | A/H3N2 | WHO CCRRI | This study | Austin Health |
| EPI_ISL_12579882 | A/Victoria/109/2022 | 3C.2a1b.2a.2 | A/H3N2 | WHO CCRRI | This study | Austin Health |
| EPI_ISL_12579883 | A/Victoria/110/2022 | 3C.2a1b.2a.2 | A/H3N2 | WHO CCRRI | This study | Austin Health |
| EPI_ISL_12579887 | A/Victoria/108/2022 | 6B.1A.5a.2 | A/H1N1 | WHO CCRRI | This study | ST Vincent Hospital |
| EPI_ISL_12579894 | A/Victoria/33/2022 | 3C.2a1b.2a.2 | A/H3N2 | WHO CCRRI | This study | ST Vincent Hospital |
| EPI_ISL_12579895 | A/Victoria/27/2022 | 3C.2a1b.2a.2 | A/H3N2 | WHO CCRRI | This study | Australian Clinical Laboratories |
| EPI_ISL_12579896 | A/Victoria/68/2022 | 3C.2a1b.2a.2 | A/H3N2 | WHO CCRRI | This study | Royal Melbourne Hospital |
| EPI_ISL_12579897 | A/Victoria/26/2022 | 3C.2a1b.2a.2 | A/H3N2 | WHO CCRRI | This study | Australian Clinical Laboratories |
| EPI_ISL_12579898 | A/Victoria/32/2022 | 3C.2a1b.2a.2 | A/H3N2 | WHO CCRRI | This study | ST Vincent Hospital |
| EPI_ISL_12579899 | A/Victoria/14/2022 | 3C.2a1b.2a.2 | A/H3N2 | WHO CCRRI | This study | Monash Medical Centre |
| EPI_ISL_12579902 | A/Victoria/78/2022 | 3C.2a1b.2a.2 | A/H3N2 | WHO CCRRI | This study | Austin Health |
| EPI_ISL_12579903 | A/Victoria/105/2022 | 3C.2a1b.2a.2 | A/H3N2 | WHO CCRRI | This study | ST Vincent Hospital |
| EPI_ISL_12579905 | A/Victoria/107/2022 | 3C.2a1b.2a.2 | A/H3N2 | WHO CCRRI | This study | ST Vincent Hospital |
| EPI_ISL_12579908 | A/Victoria/119/2022 | 3C.2a1b.2a.2 | A/H3N2 | WHO CCRRI | This study | ST Vincent Hospital |
| EPI_ISL_12579909 | A/Victoria/85/2022 | 3C.2a1b.2a.2 | A/H3N2 | WHO CCRRI | This study | Monash Medical Centre |
| EPI_ISL_12579910 | A/Victoria/120/2022 | 3C.2a1b.2a.2 | A/H3N2 | WHO CCRRI | This study | Austin Health |
| EPI_ISL_12579911 | A/Victoria/121/2022 | 3C.2a1b.2a.2 | A/H3N2 | WHO CCRRI | This study | Austin Health |
| EPI_ISL_12579913 | A/Victoria/130/2022 | 3C.2a1b.2a.2 | A/H3N2 | WHO CCRRI | This study | ST Vincent Hospital |

| | | | | | | |
|---|---|---|---|---|---|---|
| EPI_ISL_12579914 | A/Victoria/59/2022 | 3C.2a1b.2a.2 | A/H3N2 | WHO CCRRI | This study | Royal Chidrens Hospital |
| EPI_ISL_12579915 | A/Victoria/44/2022 | 3C.2a1b.2a.2 | A/H3N2 | WHO CCRRI | This study | Monash Medical Centre |
| EPI_ISL_12579916 | A/Victoria/64/2022 | 3C.2a1b.2a.2 | A/H3N2 | WHO CCRRI | This study | Monash Medical Centre |
| EPI_ISL_12579917 | A/Victoria/63/2022 | 3C.2a1b.2a.2 | A/H3N2 | WHO CCRRI | This study | Monash Medical Centre |
| EPI_ISL_12579918 | A/Victoria/73/2022 | 6B.1A.5a.2 | A/H1N1 | WHO CCRRI | This study | Alfred Hospital |
| EPI_ISL_12579919 | A/Victoria/41/2022 | 3C.2a1b.2a.2 | A/H3N2 | WHO CCRRI | This study | Alfred Hospital |
| EPI_ISL_12579920 | A/Victoria/43/2022 | 3C.2a1b.1a | A/H3N2 | WHO CCRRI | This study | Monash Medical Centre |
| EPI_ISL_12579921 | A/Victoria/125/2022 | 3C.2a1b.2a.2 | A/H3N2 | WHO CCRRI | This study | Royal Melbourne Hospital |
| EPI_ISL_12579922 | A/Victoria/126/2022 | 3C.2a1b.2a.2 | A/H3N2 | WHO CCRRI | This study | Royal Melbourne Hospital |
| EPI_ISL_12579923 | A/Victoria/79/2022 | 3C.2a1b.2a.2 | A/H3N2 | WHO CCRRI | This study | Royal Melbourne Hospital |
| EPI_ISL_12579924 | A/Victoria/101/2022 | 3C.2a1b.2a.2 | A/H3N2 | WHO CCRRI | This study | WHO CCRRI |
| EPI_ISL_12579925 | A/Victoria/76/2022 | 3C.2a1b.2a.2 | A/H3N2 | WHO CCRRI | This study | Monash Medical Centre |
| EPI_ISL_12579926 | A/Victoria/22/2022 | 3C.2a1b.2a.2 | A/H3N2 | WHO CCRRI | This study | Royal Melbourne Hospital |
| EPI_ISL_12579931 | A/Victoria/72/2022 | 3C.2a1b.2a.2 | A/H3N2 | WHO CCRRI | This study | Alfred Hospital |
| EPI_ISL_12579932 | A/Victoria/71/2022 | 3C.2a1b.2a.2 | A/H3N2 | WHO CCRRI | This study | Alfred Hospital |
| EPI_ISL_12579933 | A/Victoria/69/2022 | 3C.2a1b.2a.2 | A/H3N2 | WHO CCRRI | This study | Alfred Hospital |
| EPI_ISL_12579934 | A/Victoria/74/2022 | 3C.2a1b.2a.2 | A/H3N2 | WHO CCRRI | This study | Alfred Hospital |
| EPI_ISL_12579935 | A/Victoria/39/2022 | 3C.2a1b.2a.2 | A/H3N2 | WHO CCRRI | This study | Alfred Hospital |
| EPI_ISL_12579936 | A/Victoria/20/2022 | 3C.2a1b.2a.2 | A/H3N2 | WHO CCRRI | This study | Royal Melbourne Hospital |
| EPI_ISL_12579937 | A/Victoria/23/2022 | 3C.2a1b.2a.2 | A/H3N2 | WHO CCRRI | This study | Royal Melbourne Hospital |
| EPI_ISL_12579938 | A/Victoria/58/2022 | 3C.2a1b.2a.2 | A/H3N2 | WHO CCRRI | This study | Monash Medical Centre |
| EPI_ISL_12579939 | A/Victoria/57/2022 | 3C.2a1b.2a.2 | A/H3N2 | WHO CCRRI | This study | WHO CCRRI |
| EPI_ISL_12579942 | A/Victoria/102/2022 | 3C.2a1b.2a.2 | A/H3N2 | WHO CCRRI | This study | WHO CCRRI |
| EPI_ISL_12579943 | A/Victoria/77/2022 | 3C.2a1b.2a.2 | A/H3N2 | WHO CCRRI | This study | ST Vincent Hospital |
| EPI_ISL_12579944 | A/Victoria/115/2022 | 3C.2a1b.2a.2 | A/H3N2 | WHO CCRRI | This study | Monash Medical Centre |
| EPI_ISL_12579945 | A/Victoria/83/2022 | 6B.1A.5a.2 | A/H1N1 | WHO CCRRI | This study | Monash Medical Centre |
| EPI_ISL_12579946 | A/Victoria/81/2022 | 3C.2a1b.2a.2 | A/H3N2 | WHO CCRRI | This study | Monash Medical Centre |
| EPI_ISL_12579947 | A/Victoria/80/2022 | 3C.2a1b.2a.2 | A/H3N2 | WHO CCRRI | This study | Monash Medical Centre |
| EPI_ISL_12579948 | A/Victoria/87/2022 | 3C.2a1b.2a.2 | A/H3N2 | WHO CCRRI | This study | Monash Medical Centre |
| EPI_ISL_12579949 | A/Victoria/84/2022 | 3C.2a1b.2a.2 | A/H3N2 | WHO CCRRI | This study | Monash Medical Centre |
| EPI_ISL_12579950 | A/Victoria/86/2022 | 3C.2a1b.2a.2 | A/H3N2 | WHO CCRRI | This study | Monash Medical Centre |
| EPI_ISL_12579951 | A/Victoria/37/2022 | 3C.2a1b.2a.2 | A/H3N2 | WHO CCRRI | This study | Alfred Hospital |
| EPI_ISL_12579952 | A/Victoria/113/2022 | 3C.2a1b.2a.2 | A/H3N2 | WHO CCRRI | This study | Monash Medical Centre |
| EPI_ISL_12579953 | A/Victoria/111/2022 | 3C.2a1b.2a.2 | A/H3N2 | WHO CCRRI | This study | Monash Medical Centre |
| EPI_ISL_12579954 | A/Victoria/112/2022 | 3C.2a1b.2a.2 | A/H3N2 | WHO CCRRI | This study | Monash Medical Centre |
| EPI_ISL_12579956 | A/Victoria/70/2022 | 3C.2a1b.2a.2 | A/H3N2 | WHO CCRRI | This study | Alfred Hospital |
| EPI_ISL_12850597 | A/Victoria/60/2022 | 6B.1A.5a.2 | A/H1N1 | WHO CCRRI | This study | WHO CCRRI |
| EPI_ISL_12850600 | A/Victoria/412/2022 | 6B.1A.5a.2 | A/H1N1 | WHO CCRRI | This study | Royal Melbourne Hospital |
| EPI_ISL_12850879 | A/Victoria/341/2022 | 3C.2a1b.2a.2 | A/H3N2 | WHO CCRRI | This study | ST Vincent Hospital |
| EPI_ISL_12850890 | A/Victoria/1152/2022 | 3C.2a1b.2a.2 | A/H3N2 | WHO CCRRI | This study | WHO CCRRI |
| EPI_ISL_12850901 | A/Victoria/371/2022 | 3C.2a1b.2a.2 | A/H3N2 | WHO CCRRI | This study | WHO CCRRI |
| EPI_ISL_12850911 | A/Victoria/693/2022 | 3C.2a1b.2a.2 | A/H3N2 | WHO CCRRI | This study | Austin Health |
| EPI_ISL_12850922 | A/Victoria/209/2022 | 3C.2a1b.2a.2 | A/H3N2 | WHO CCRRI | This study | Alfred Hospital |
| EPI_ISL_12850932 | A/Victoria/232/2022 | 3C.2a1b.2a.2 | A/H3N2 | WHO CCRRI | This study | Alfred Hospital |
| EPI_ISL_12850942 | A/Victoria/234/2022 | 3C.2a1b.2a.2 | A/H3N2 | WHO CCRRI | This study | Alfred Hospital |
| EPI_ISL_12850952 | A/Victoria/273/2022 | 3C.2a1b.2a.2 | A/H3N2 | WHO CCRRI | This study | Alfred Hospital |
| EPI_ISL_12851004 | A/Victoria/254/2022 | 3C.2a1b.2a.2 | A/H3N2 | WHO CCRRI | This study | Royal Melbourne Hospital |
| EPI_ISL_12851014 | A/Victoria/410/2022 | 3C.2a1b.2a.2 | A/H3N2 | WHO CCRRI | This study | Royal Melbourne Hospital |
| EPI_ISL_12851024 | A/Victoria/391/2022 | 3C.2a1b.2a.2 | A/H3N2 | WHO CCRRI | This study | Royal Melbourne Hospital |
| EPI_ISL_12851034 | A/Victoria/437/2022 | 3C.2a1b.2a.2 | A/H3N2 | WHO CCRRI | This study | Royal Melbourne Hospital |
| EPI_ISL_12851045 | A/Victoria/266/2022 | 3C.2a1b.2a.2 | A/H3N2 | WHO CCRRI | This study | Alfred Hospital |
| EPI_ISL_12851076 | A/Victoria/278/2022 | 3C.2a1b.2a.2 | A/H3N2 | WHO CCRRI | This study | Alfred Hospital |
| EPI_ISL_12851108 | A/Victoria/315/2022 | 3C.2a1b.2a.2 | A/H3N2 | WHO CCRRI | This study | Monash Medical Centre |
| EPI_ISL_12851109 | A/Victoria/310/2022 | 3C.2a1b.2a.2 | A/H3N2 | WHO CCRRI | This study | Monash Medical Centre |
| EPI_ISL_12851110 | A/Victoria/311/2022 | 3C.2a1b.2a.2 | A/H3N2 | WHO CCRRI | This study | Monash Medical Centre |

| | | | | | | |
|---|---|---|---|---|---|---|
| EPI_ISL_12851111 | A/Victoria/307/2022 | 3C.2a1b.2a.2 | A/H3N2 | WHO CCRRI | This study | Monash Medical Centre |
| EPI_ISL_12851112 | A/Victoria/290/2022 | 3C.2a1b.2a.2 | A/H3N2 | WHO CCRRI | This study | Monash Medical Centre |
| EPI_ISL_12851113 | A/Victoria/298/2022 | 3C.2a1b.2a.2 | A/H3N2 | WHO CCRRI | This study | Monash Medical Centre |
| EPI_ISL_13168836 | A/Victoria/65a/2022 | 3C.2a1b.2a.2 | A/H3N2 | WHO CCRRI | This study | Royal Melbourne Hospital |
| EPI_ISL_13168837 | A/Victoria/100/2022 | 3C.2a1b.2a.2 | A/H3N2 | WHO CCRRI | This study | Austin Health |
| EPI_ISL_13168838 | A/Victoria/35/2022 | 3C.2a1b.2a.2 | A/H3N2 | WHO CCRRI | This study | Alfred Hospital |
| EPI_ISL_13168839 | A/Victoria/36/2022 | 3C.2a1b.2a.2 | A/H3N2 | WHO CCRRI | This study | Alfred Hospital |
| EPI_ISL_13168840 | A/Victoria/38/2022 | 3C.2a1b.2a.2 | A/H3N2 | WHO CCRRI | This study | Alfred Hospital |
| EPI_ISL_13168841 | A/Victoria/34/2022 | 3C.2a1b.2a.2 | A/H3N2 | WHO CCRRI | This study | Austin Health |
| EPI_ISL_13168842 | A/Victoria/75/2022 | 3C.2a1b.2a.2 | A/H3N2 | WHO CCRRI | This study | Victorian Infectious Diseases Reference Laboratory |
| EPI_ISL_13168843 | A/Victoria/21/2022 | 3C.2a1b.2a.2 | A/H3N2 | WHO CCRRI | This study | Royal Melbourne Hospital |
| EPI_ISL_13168844 | A/Victoria/48/2022 | 3C.2a1b.2a.2 | A/H3N2 | WHO CCRRI | This study | ST Vincent Hospital |
| EPI_ISL_13175618 | A/Victoria/984/2022 | 3C.2a1b.2a.2 | A/H3N2 | WHO CCRRI | This study | Melbourne Pathology |
| EPI_ISL_13175622 | A/Victoria/993/2022 | 3C.2a1b.2a.2 | A/H3N2 | WHO CCRRI | This study | Melbourne Pathology |
| EPI_ISL_13175639 | A/Victoria/811/2022 | 3C.2a1b.2a.2 | A/H3N2 | WHO CCRRI | This study | Austin Health |
| EPI_ISL_13175641 | A/Victoria/810/2022 | 3C.2a1b.2a.2 | A/H3N2 | WHO CCRRI | This study | Austin Health |
| EPI_ISL_13175642 | A/Victoria/818/2022 | 3C.2a1b.2a.2 | A/H3N2 | WHO CCRRI | This study | Austin Health |
| EPI_ISL_13175651 | A/Victoria/1023/2022 | 3C.2a1b.2a.2 | A/H3N2 | WHO CCRRI | This study | Melbourne Pathology |
| EPI_ISL_13175656 | A/Victoria/1007/2022 | 3C.2a1b.2a.2 | A/H3N2 | WHO CCRRI | This study | Melbourne Pathology |
| EPI_ISL_13398668 | A/Victoria/221/2022 | 6B.1A.5a.2 | A/H1N1 | WHO CCRRI | This study | Alfred Hospital |
| EPI_ISL_13398682 | A/Victoria/883/2022 | 3C.2a1b.2a.2 | A/H3N2 | WHO CCRRI | This study | Monash Medical Centre |
| EPI_ISL_13398683 | A/Victoria/863/2022 | 3C.2a1b.2a.2 | A/H3N2 | WHO CCRRI | This study | Monash Medical Centre |
| EPI_ISL_13398684 | A/Victoria/856/2022 | 3C.2a1b.2a.2 | A/H3N2 | WHO CCRRI | This study | Monash Medical Centre |
| EPI_ISL_13398685 | A/Victoria/858/2022 | 3C.2a1b.2a.2 | A/H3N2 | WHO CCRRI | This study | Monash Medical Centre |
| EPI_ISL_13398686 | A/Victoria/851/2022 | 3C.2a1b.2a.2 | A/H3N2 | WHO CCRRI | This study | Monash Medical Centre |
| EPI_ISL_13531020 | A/Victoria/718/2022 | 3C.2a1b.2a.2 | A/H3N2 | WHO CCRRI | This study | Monash Medical Centre |
| EPI_ISL_13531021 | A/Victoria/723/2022 | 3C.2a1b.2a.2 | A/H3N2 | WHO CCRRI | This study | Monash Medical Centre |
| EPI_ISL_13531022 | A/Victoria/698/2022 | 3C.2a1b.2a.2 | A/H3N2 | WHO CCRRI | This study | Monash Medical Centre |
| EPI_ISL_13531023 | A/Victoria/708/2022 | 3C.2a1b.2a.2 | A/H3N2 | WHO CCRRI | This study | Monash Medical Centre |
| EPI_ISL_13531024 | A/Victoria/785/2022 | 3C.2a1b.2a.2 | A/H3N2 | WHO CCRRI | This study | Monash Medical Centre |
| EPI_ISL_13531181 | A/Victoria/871/2022 | 3C.2a1b.2a.2 | A/H3N2 | WHO CCRRI | This study | Monash Medical Centre |
| EPI_ISL_13531184 | A/Victoria/848/2022 | 3C.2a1b.2a.2 | A/H3N2 | WHO CCRRI | This study | Monash Medical Centre |
| EPI_ISL_13531226 | A/Victoria/757/2022 | 3C.2a1b.2a.2 | A/H3N2 | WHO CCRRI | This study | Monash Medical Centre |
| EPI_ISL_13531227 | A/Victoria/864/2022 | 3C.2a1b.2a.2 | A/H3N2 | WHO CCRRI | This study | Monash Medical Centre |
| EPI_ISL_13531228 | A/Victoria/98/2022 | 3C.2a1b.2a.2 | A/H3N2 | WHO CCRRI | This study | Austin Health |
| EPI_ISL_13531233 | A/Victoria/174/2022 | 3C.2a1b.2a.2 | A/H3N2 | WHO CCRRI | This study | Melbourne Pathology |
| EPI_ISL_13531234 | A/Victoria/170/2022 | 3C.2a1b.2a.2 | A/H3N2 | WHO CCRRI | This study | Melbourne Pathology |
| EPI_ISL_13531235 | A/Victoria/172/2022 | 3C.2a1b.2a.2 | A/H3N2 | WHO CCRRI | This study | Melbourne Pathology |
| EPI_ISL_13531236 | A/Victoria/169/2022 | 3C.2a1b.2a.2 | A/H3N2 | WHO CCRRI | This study | Melbourne Pathology |
| EPI_ISL_13655099 | A/Victoria/139/2022 | 3C.2a1b.2a.2 | A/H3N2 | WHO CCRRI | This study | Monash Medical Centre |
| EPI_ISL_13655101 | A/Victoria/138/2022 | 3C.2a1b.2a.2 | A/H3N2 | WHO CCRRI | This study | Monash Medical Centre |
| EPI_ISL_13655102 | A/Victoria/158/2022 | 3C.2a1b.2a.2 | A/H3N2 | WHO CCRRI | This study | Monash Medical Centre |
| EPI_ISL_13655105 | A/Victoria/404/2022 | 3C.2a1b.2a.2 | A/H3N2 | WHO CCRRI | This study | Alfred Hospital |
| EPI_ISL_13655108 | A/Victoria/519/2022 | 3C.2a1b.2a.2 | A/H3N2 | WHO CCRRI | This study | Alfred Hospital |
| EPI_ISL_13655109 | A/Victoria/521/2022 | 3C.2a1b.2a.2 | A/H3N2 | WHO CCRRI | This study | Alfred Hospital |
| EPI_ISL_13655114 | A/Victoria/775/2022 | 3C.2a1b.2a.2 | A/H3N2 | WHO CCRRI | This study | Monash Medical Centre |
| EPI_ISL_13655115 | A/Victoria/704/2022 | 3C.2a1b.2a.2 | A/H3N2 | WHO CCRRI | This study | Monash Medical Centre |
| EPI_ISL_13983918 | A/Victoria/747/2022 | 6B.1A.5a.2 | A/H1N1 | WHO CCRRI | This study | Monash Medical Centre |
| EPI_ISL_13983919 | A/Victoria/779/2022 | 6B.1A.5a.2 | A/H1N1 | WHO CCRRI | This study | Monash Medical Centre |
| EPI_ISL_13983978 | A/Victoria/532/2022 | 3C.2a1b.2a.2 | A/H3N2 | WHO CCRRI | This study | Royal Melbourne Hospital |
| EPI_ISL_13983982 | A/Victoria/936/2022 | 3C.2a1b.2a.2 | A/H3N2 | WHO CCRRI | This study | Alfred Hospital |
| EPI_ISL_13984023 | A/Victoria/938/2022 | 3C.2a1b.2a.2 | A/H3N2 | WHO CCRRI | This study | Alfred Hospital |
| EPI_ISL_14115533 | A/Victoria/143/2022 | 3C.2a1b.2a.2 | A/H3N2 | WHO CCRRI | This study | Royal Melbourne Hospital |
| EPI_ISL_14161750 | A/Victoria/783/2022 | 3C.2a1b.2a.2 | A/H3N2 | WHO CCRRI | This study | Monash Medical Centre |

| | | | | | | |
|---|---|---|---|---|---|---|
| EPI_ISL_14161752 | A/Victoria/758/2022 | 3C.2a1b.2a.2 | A/H3N2 | WHO CCRRI | This study | Monash Medical Centre |
| EPI_ISL_14161754 | A/Victoria/789/2022 | 3C.2a1b.2a.2 | A/H3N2 | WHO CCRRI | This study | Monash Medical Centre |
| EPI_ISL_14161756 | A/Victoria/844/2022 | 3C.2a1b.2a.2 | A/H3N2 | WHO CCRRI | This study | Monash Medical Centre |
| EPI_ISL_14161758 | A/Victoria/855/2022 | 3C.2a1b.2a.2 | A/H3N2 | WHO CCRRI | This study | Monash Medical Centre |
| EPI_ISL_14161760 | A/Victoria/878/2022 | 3C.2a1b.2a.2 | A/H3N2 | WHO CCRRI | This study | Monash Medical Centre |
| EPI_ISL_14161764 | A/Victoria/519/2022 | 3C.2a1b.2a.2 | A/H3N2 | WHO CCRRI | This study | Alfred Hospital |
| EPI_ISL_14161766 | A/Victoria/521/2022 | 3C.2a1b.2a.2 | A/H3N2 | WHO CCRRI | This study | Alfred Hospital |
| EPI_ISL_14161794 | A/Victoria/82/2022 | 3C.2a1b.2a.2 | A/H3N2 | WHO CCRRI | This study | Monash Medical Centre |
| EPI_ISL_14161797 | A/Victoria/88/2022 | 3C.2a1b.2a.2 | A/H3N2 | WHO CCRRI | This study | Monash Medical Centre |
| EPI_ISL_14161799 | A/Victoria/104/2022 | 3C.2a1b.2a.2 | A/H3N2 | WHO CCRRI | This study | Royal Melbourne Hospital |
| EPI_ISL_14161802 | A/Victoria/210/2022 | 3C.2a1b.2a.2 | A/H3N2 | WHO CCRRI | This study | Alfred Hospital |
| EPI_ISL_14161805 | A/Victoria/213/2022 | 3C.2a1b.2a.2 | A/H3N2 | WHO CCRRI | This study | Alfred Hospital |
| EPI_ISL_14161808 | A/Victoria/212/2022 | 3C.2a1b.2a.2 | A/H3N2 | WHO CCRRI | This study | Alfred Hospital |
| EPI_ISL_14161811 | A/Victoria/211/2022 | 3C.2a1b.2a.2 | A/H3N2 | WHO CCRRI | This study | Alfred Hospital |
| EPI_ISL_14161813 | A/Victoria/214/2022 | 3C.2a1b.2a.2 | A/H3N2 | WHO CCRRI | This study | Alfred Hospital |
| EPI_ISL_14161816 | A/Victoria/215/2022 | 3C.2a1b.2a.2 | A/H3N2 | WHO CCRRI | This study | Alfred Hospital |
| EPI_ISL_14161819 | A/Victoria/216/2022 | 3C.2a1b.2a.2 | A/H3N2 | WHO CCRRI | This study | Alfred Hospital |
| EPI_ISL_14161821 | A/Victoria/508/2022 | 3C.2a1b.2a.2 | A/H3N2 | WHO CCRRI | This study | Alfred Hospital |
| EPI_ISL_14161833 | A/Victoria/760/2022 | 3C.2a1b.2a.2 | A/H3N2 | WHO CCRRI | This study | Monash Medical Centre |
| EPI_ISL_14189363 | A/Victoria/772/2022 | 3C.2a1b.2a.2 | A/H3N2 | WHO CCRRI | This study | Monash Medical Centre |
| EPI_ISL_14467656 | A/Newcastle/47/2022 | 3C.2a1b.2a.2 | A/H3N2 | WHO CCRRI | This study | John Hunter Hospital |
| EPI_ISL_14819723 | A/Victoria/220/2022 | 3C.2a1b.2a.2 | A/H3N2 | WHO CCRRI | This study | Alfred Hospital |
| EPI_ISL_14819724 | A/Victoria/231/2022 | 3C.2a1b.2a.2 | A/H3N2 | WHO CCRRI | This study | Alfred Hospital |
| EPI_ISL_15833316 | A/Victoria/1414/2022 | 3C.2a1b.2a.2 | A/H3N2 | WHO CCRRI | This study | Monash Medical Centre |
| EPI_ISL_15833317 | A/Victoria/1382/2022 | 3C.2a1b.2a.2 | A/H3N2 | WHO CCRRI | This study | Monash Medical Centre |
| EPI_ISL_15833389 | A/Victoria/1406/2022 | 3C.2a1b.2a.2 | A/H3N2 | WHO CCRRI | This study | Monash Medical Centre |
| EPI_ISL_15833411 | A/Victoria/2609/2022 | 3C.2a1b.2a.2 | A/H3N2 | WHO CCRRI | This study | Monash Medical Centre |
| EPI_ISL_9673931 | A/Victoria/7/2021 | 3C.2a1b.2a.2 | A/H3N2 | WHO CCRRI | This study | Monash Medical Centre |
| EPI_ISL_9673932 | A/Victoria/9/2021 | 3C.2a1b.2a.2 | A/H3N2 | WHO CCRRI | This study | Alfred Hospital |
| EPI_ISL_9847985 | A/Victoria/11/2021 | 3C.2a1b.2a.2 | A/H3N2 | WHO CCRRI | This study | Royal Melbourne Hospital |
| EPI_ISL_9847986 | A/Victoria/16/2021 | 3C.2a1b.2a.2 | A/H3N2 | WHO CCRRI | This study | Royal Melbourne Hospital |
| EPI_ISL_9847988 | A/Victoria/7/2021 | 3C.2a1b.2a.2 | A/H3N2 | WHO CCRRI | This study | Monash Medical Centre |

[a]World Health Organisation for Reference and Research on Influenza (WHO CCRRI)

Supplementary Table 4. GISAID Accession numbers for international and reference influenza viruses displayed in the phylogenetic tree (Figure 3)

| Isolate_Id | Isolate_Name | Submitting_Lab | Authors | Originating_Lab |
|---|---|---|---|---|
| EPI_ISL_13044336 | A/Tumbes/FPT02314/2022(H3N2) | NAMRU-6[a] | Gilda Troncos, Victoria Espejo, Cristhopher Cruz, Victor Herrera Sunci?n , Yeny Tinoco, Sonia Ampuero, Marita Silva, Paul Graf. | NAMRU-6 |
| EPI_ISL_13044334 | A/Tumbes/FPT02308/2022(H3N2) | NAMRU-6 | Gilda Troncos, Victoria Espejo, Cristhopher Cruz, Victor Herrera Sunci?n , Yeny Tinoco, Sonia Ampuero, Marita Silva, Paul Graf. | NAMRU-6 |
| EPI_ISL_11171190 | A/Tumbes/FPT02244/2021(H3N2) | NAMRU-6 | Gilda Troncos, Victoria Espejo, Cristhopher Cruz, Victor Herrera Sunci?n , Yeny Tinoco, Sonia Ampuero, Marita Silva, Paul Graf. | NAMRU-6 |
| EPI_ISL_11171179 | A/Bagua/FPG00828/2022(H3N2) | NAMRU-6 | Gilda Troncos, Victoria Espejo, Cristhopher Cruz, Jorge Miguel Alexander Olivos Villanueva, Ronald Eduardo Ramos Neyra , Yeny Tinoco, Sonia Ampuero, Marita Silva, Paul Graf. | NAMRU-6 |
| EPI_ISL_11400612 | A/Michigan/UOM10043998503/2022 | University of Michigan | | University of Michigan Clinical Microbiology Laboratory |
| EPI_ISL_6902173 | A/Michigan/UOM10042604678/2021 | University of Michigan | | University of Michigan Clinical Microbiology Laboratory |
| EPI_ISL_13148944 | A/Human/New York/PV60633/2022 | Icahn School of Medicine at Mount Sinai | Gonzalez-Reiche, Ana S.; Alshammary, Hala; Polanco, Jose; Kim, Eunhye; Amoako, Angela; Rooker, Aria; Cognigni, Christian; Floda, Daniel; van de Guchte, Adriana; Khalil, Zain; Srivastava, Komal; Sebra, Robert; Garc?a-Sastre, Adolfo; Patel, Gopi; Schaefer, Sarah; Ramirez, Juan David; Banu, Radhika; Shrestha, Paras; Paniz-Mondolfi, Alberto; Sordillo, Emilia Mia; Simon, Viviana; van Bakel, Harm | Icahn School of Medicine at Mount Sinai |
| EPI_ISL_8769986 | A/Denmark/17/2021 | Statens Serum Institute | Trebbien, Ramona; Nissen, Jakob; Bolt Botnen, Amanda | Statens Serum Institute |
| EPI_ISL_8694331 | A/Denmark/39/2021 | Statens Serum Institute | Trebbien, Ramona; Nissen, Jakob; Bolt Botnen, Amanda | Statens Serum Institute |
| EPI_ISL_12323526 | A/Denmark/288/2022 | Statens Serum Institute | Trebbien, Ramona; Nissen, Jakob; Bolt Botnen, Amanda | Statens Serum Institute |
| EPI_ISL_13326477 | A/Austria/MUW_1510488/2022 | Medical University of Vienna | Camp, J.V.; Redlberger-Fritz, M. | Department of Virology, Medical University Vienna |
| EPI_ISL_11403058 | A/Lisboa/23/2022 | Instituto Nacional de Saúde Dr. Ricardo Jorge | Guiomar, R.; Melo, A.; Costa, I.; Conde, P, Henriques, C. | Instituto Nacional de Saúde Dr. Ricardo Jorge |
| EPI_ISL_9775384 | A/Luxembourg/LNS6784733/2021 | Laboratoire National de Santé | Anke Wienecke-Baldacchino, Sibel Berger, Guillaume Fournier | Laboratoire National de Santé |
| EPI_ISL_13431097 | A/England/220840504/2022 | UK Health Security Agency - Colindale | UKHSA, Respiratory Virus Unit | UK Health Security Agency - Colindale |
| EPI_ISL_9383724 | A/England/215121919/2021 | UK Health Security Agency - Colindale | UKHSA, Respiratory Virus Unit | UK Health Security Agency - Colindale |
| EPI_ISL_5935472 | A/England/214100500/2021 | UK Health Security Agency - Colindale | | UK Health Security Agency - Colindale |
| EPI_ISL_7607226 | A/Netherlands/10148/2021 | National Institute for Public Health and the Environment (RIVM) | A,Meijer; S, vd Brink; L,Wijsman | PAMM |
| EPI_ISL_12064265 | A/Netherlands/10905/2022 | National Institute for Public Health and the Environment (RIVM) | A,Meijer; L,Wijsman; S, vd Brink | Diakonessenhuis Utrecht |
| EPI_ISL_9627675 | A/Israel/S-1/2021 | Crick Worldwide Influenza Centre | | Central Virology Laboratory Israel (NIC) |
| EPI_ISL_9533449 | A/Israel/R-724/2022 | Crick Worldwide Influenza Centre | | Central Virology Laboratory Israel (NIC) |
| EPI_ISL_9842667 | A/Norway/521/2022 | Crick Worldwide Influenza Centre | | WHO National Influenza Centre |
| EPI_ISL_12942424 | A/Ghana/195/2022 | Crick Worldwide Influenza Centre | | University of Ghana |
| EPI_ISL_9593247 | A/Iran/15/Azar21/2021 | Crick Worldwide Influenza Centre | | Tehran University of Medical Sciences |
| EPI_ISL_9593240 | A/Iran/83/Ab/8A/2021 | Crick Worldwide Influenza Centre | | Tehran University of Medical Sciences |
| EPI_ISL_12219314 | A/Serbia/4/2021 | Crick Worldwide Influenza Centre | | Institute of Immunology and Virology Torlak |
| EPI_ISL_13017096 | A/Morocco/82/2021 | Crick Worldwide Influenza Centre | | Institut National d'Hygiène |
| EPI_ISL_13339904 | A/Ireland/17354/2022 | Crick Worldwide Influenza Centre | | UCD National Virus Reference Laboratory |
| EPI_ISL_9842672 | A/Ireland/2795/2022 | Crick Worldwide Influenza Centre | | UCD National Virus Reference Laboratory |

| | | | | |
|---|---|---|---|---|
| EPI_ISL_8808670 | A/Ireland/92285/2021 | Crick Worldwide Influenza Centre | | UCD National Virus Reference Laboratory |
| EPI_ISL_8808663 | A/Ireland/82/2021 | Crick Worldwide Influenza Centre | | UCD National Virus Reference Laboratory |
| EPI_ISL_13203500 | A/Iceland/28760/2022 | Crick Worldwide Influenza Centre | | Landspitali - University Hospital |
| EPI_ISL_13203460 | A/Iceland/33333/2022 | Crick Worldwide Influenza Centre | | Landspitali - University Hospital |
| EPI_ISL_9627689 | A/Romania/495025/2021 | Crick Worldwide Influenza Centre | | Cantacuzino Institute |
| EPI_ISL_9593243 | A/Hungary/8/2021 | Crick Worldwide Influenza Centre | | Central Veterinary Institute Budapest |
| EPI_ISL_9593233 | A/Hungary/2/2022 | Crick Worldwide Influenza Centre | | Central Veterinary Institute Budapest |
| EPI_ISL_12219613 | A/Bosnia and Herzegovina/103/2021 | Crick Worldwide Influenza Centre | | University of Sarajevo |
| EPI_ISL_12219468 | A/Cote d'Ivoire/4032/2021 | Crick Worldwide Influenza Centre | | Pasteur Institut of Côte d'Ivoire |
| EPI_ISL_12219451 | A/Cote d'Ivoire/4120/2021 | Crick Worldwide Influenza Centre | | Pasteur Institut of Côte d'Ivoire |
| EPI_ISL_12219432 | A/Cote d'Ivoire/8/2022 | Crick Worldwide Influenza Centre | | Pasteur Institut of Côte d'Ivoire |
| EPI_ISL_12219308 | A/Kosova/954/2022 | Crick Worldwide Influenza Centre | | National Institute of Public Health of Kosova |
| EPI_ISL_12240159 | A/Netherlands/10024/2021 | Crick Worldwide Influenza Centre | | National Institute for Public Health and the Environment (RIVM) |
| EPI_ISL_13203484 | A/Switzerland/83625/2022 | Crick Worldwide Influenza Centre | | Swiss National Reference Centre for Influenza |
| EPI_ISL_9401999 | A/Switzerland/69954/2021 | Crick Worldwide Influenza Centre | | Swiss National Reference Centre for Influenza |
| EPI_ISL_12839635 | A/Sachsen/7/2022 | Robert Koch Institute Nationales Referenzzentrum für Influenza | | Robert Koch-Institute Nationales Referenzzentrum für Influenza |
| EPI_ISL_11217722 | A/Hessen/4/2022 | Robert Koch Institute Nationales Referenzzentrum für Influenza | | Robert Koch-Institute Nationales Referenzzentrum für Influenza |
| EPI_ISL_13812784 | A/United Kingdom/13176/2022 | U.S. Air Force School of Aerospace Medicine | Gruner, W.E., Muehleman, D.M., Fries, A.C., Garrett, C.M., Hanson, J.F., DeMarcus, L.S., Sjoberg, P.A., Wasik, P.M., Robbins, A.S., Macias, E.A., Shaw, K.B., Selent, M.U. | U.S. Air Force School of Aerospace Medicine |
| EPI_ISL_12422440 | A/Texas/12644/2021 | U.S. Air Force School of Aerospace Medicine | Gruner, W.E., Muehleman, D.M., Fries, A.C., Garrett, C.M., Hanson, J.F., DeMarcus, L.S., Sjoberg, P.A., Wasik, P.M., Robbins, A.S., Macias, E.A., Shaw, K.B., Selent, M.U. | U.S. Air Force School of Aerospace Medicine |
| EPI_ISL_7404317 | A/Maryland/12435/2021 | U.S. Air Force School of Aerospace Medicine | Gruner, W.E., Muehleman, D.M., Fries, A.C., Garrett, C.M., Hanson, J.F., DeMarcus, L.S., Sjoberg, P.A., Wasik, P.M., Robbins, A.S., Macias, E.A., Shaw, K.B., Selent, M.U. | U.S. Air Force School of Aerospace Medicine |
| EPI_ISL_9994834 | A/Catalonia/NSVH101718786/2022 | Hospital Universitari Vall d'Hebron | Anton, A.; Martin, MC.; Andres, C.; Codina, MG.; Pumarola, T. | Hospital Universitari Vall d'Hebron |
| EPI_ISL_12609685 | A/Romania/509797_BB/2022 | Cantacuzino Institute | Luiza,Ustea; Oana,Vitencu; Catalina,Pascu; Sorin,Dinu; Mihaela,Lazar | Cantacuzino Institute |
| EPI_ISL_12607361 | A/Romania/510965_GR/2022 | Cantacuzino Institute | Luiza,Ustea; Oana,Vitencu; Catalina,Pascu; Sorin,Dinu; Mihaela,Lazar | Cantacuzino Institute |
| EPI_ISL_12646189 | A/Andalucia/1528/2022 | Instituto de Salud Carlos III | Pozo,F; Iglesias-Caballero, M.; Gonz?lez-Esguevillas,M.; Molinero,M.; Camarero,S.; Mart?n,V.; Reyes,N.; Sandon?s,V.; V?zquez-Mor?n,S.; Casas,I. | Servicio de Microbiología Hospital Virgen de las Nieves |
| EPI_ISL_10337732 | A/Netherlands/00090/2021 | Erasmus Medical Center | | Erasmus Medical Center |
| EPI_ISL_9760141 | A/Netherlands/00048/2022 | Erasmus Medical Center | | Erasmus Medical Center |
| EPI_ISL_14033333 | A/Guizhou-Anlong/319/2022 | WHO Chinese National Influenza Center | Zeng xiaoxu, Li Xiyan, Huang Weijuan, Dayan Wang | WHO Chinese National Influenza Center |
| EPI_ISL_14033290 | A/Guizhou-Liuzhite/326/2022 | WHO Chinese National Influenza Center | Zeng xiaoxu, Li Xiyan, Huang Weijuan, Dayan Wang | WHO Chinese National Influenza Center |
| EPI_ISL_14033215 | A/Guangxi-Fangcheng/11331/2021 | WHO Chinese National Influenza Center | Zeng xiaoxu, Li Xiyan, Huang Weijuan, Dayan Wang | WHO Chinese National Influenza Center |
| EPI_ISL_13440481 | A/YOKOHAMA/3/2021 | National Institute of Infectious Diseases | Takashita,Emi;Fujisaki,Seiichiro;Watanabe,Shinji;Hasegawa,Hideki | Yokohama City Institute of Public Health. |
| EPI_ISL_13284206 | A/Colombia/2496/2022 | CDC USA[b] | | Instituto Nacional de Salud de Columbia |
| EPI_ISL_13580811 | A/El Salvador/9212/2022 | CDC USA | | Contiguo a Hospital Rosales |
| EPI_ISL_12710455 | A/Madagascar/3048/2021 | CDC USA | | Institut Pasteur de Madagascar |
| EPI_ISL_13892666 | A/Serbia/2189/2022 | CDC USA | | Institute of Immunology and Virology Torlak |

| | | | | |
|---|---|---|---|---|
| EPI_ISL_13892659 | A/Serbia/2262/2022 | CDC USA | | Institute of Immunology and Virology Torlak |
| EPI_ISL_8808407 | A/Abu Dhabi/3550476/2021 | CDC USA | | Shaikh Khalifa Medical City |
| EPI_ISL_13284263 | A/Argentina/2214/2022 | CDC USA | | Instituto Nacional de Enfermedades Infecciosas |
| EPI_ISL_10828452 | A/India/Pun-NIV60/2021 | CDC USA | | National Institute of Virology |
| EPI_ISL_10826393 | A/Kenya/101/2022 | CDC USA | | CDC-Kenya |
| EPI_ISL_13284264 | A/Nicaragua/2624/2022 | CDC USA | | Laboratorio de Virologia, Direccion de Microbiologia |
| EPI_ISL_9346127 | A/Pakistan/ICT12/2021 | CDC USA | | National Institute of Health |
| EPI_ISL_9346122 | A/Pakistan/KE3/2021 | CDC USA | | National Institute of Health |
| EPI_ISL_13284202 | A/Uruguay/127/2022 | CDC USA | | Departamento de Laboratorio de Salud Pública (DLSP) |
| EPI_ISL_14036331 | A/Guyane/365/2021 | CDC USA | | National Influenza Center French Guiana and French Indies |
| EPI_ISL_13343701 | A/Puerto Rico/02/2022 | CDC USA | | Puerto Rico Department of Health |
| EPI_ISL_13284051 | A/Puerto Rico/06/2022 | CDC USA | | Puerto Rico Department of Health |
| EPI_ISL_10493003 | A/Puerto Rico/04/2021 | CDC USA | | Puerto Rico Department of Health |
| EPI_ISL_10827595 | A/Minnesota/06/2022 | CDC USA | | Minnesota Department of Health |
| EPI_ISL_11969459 | A/Congo/026/2022 | CDC USA | | INRB Service de Virologie |
| EPI_ISL_9388974 | A/Ethiopia/0603/2021 | CDC USA | | Ethiopian Public Health Institute, National Influenza Laboratory |
| EPI_ISL_9389004 | A/Bangladesh/4037/2021 | CDC USA | | icddr,b International Centre for Diarrhoeal Disease Research, Bangladesh |
| EPI_ISL_13892749 | A/Dominican Republic/195/2022 | CDC USA | | Laboratorio Nacional de Salud Publica Dr. Defillo |
| EPI_ISL_13284698 | A/Niger/10289/2022 | CDC USA | | Centre de Recherche Medicale et Sanitaire (CERMES) |
| EPI_ISL_11956882 | A/Nigeria/6593/2021 | CDC USA | | NCDC Public Health Reference Laboratory |
| EPI_ISL_13655578 | A/Singapore/KK0015/2022 | Ministry of Health, Singapore | Chen,B.;Zhou, Z.; Yeo,B.; Cui,L; Lin,R.T.P. | Ministry of Health, Singapore |
| EPI_ISL_8640747 | A/Singapore/INFTT0006/2021 | Ministry of Health, Singapore | Chen,B.;Zhou,Z.; Ching,K.C.; Cui,L; Lin,R.T.P. | Ministry of Health, Singapore |
| EPI_ISL_9067303 | A/Sweden/15/2021 | Public Health Agency of Sweden | | Public Health Agency of Sweden |
| EPI_ISL_12809147 | A/Lorraine/29637/2022 | Institut Pasteur | | Institut Pasteur |
| EPI_ISL_11323853 | A/PAPEETE/51874/2021 | Institut Pasteur | | Institut Pasteur |

[a]Naval Medical Research Unit 6 (NAMRU-6)

[b]Centers for Disease Control and Prevention

Supplementary Table 5. Comparison of demographics between interviewed and non-interviewed influenza cases, Victoria, Australia, 1 November 2021 – 6 April 2022

| | Interviewed (n = 124) | | Not interviewed (n = 76) | | p-value[a] |
|---|---|---|---|---|---|
| | n | % | n | % | |
| Sex | | | | | 0.51 |
| - Female | 66 | 53.2% | 36 | 47.4% | |
| - Male | 58 | 46.8% | 40 | 52.6% | |
| Age group (years) | | | | | 0.86 |
| - <5 | 6 | 4.8% | 3 | 3.9% | |
| - 5-14 | 12 | 9.7% | 5 | 6.6% | |
| - 15-24 | 66 | 53.2% | 45 | 59.2% | |
| - 25-34 | 22 | 17.7% | 12 | 15.8% | |
| - 35-44 | 1 | 0.8% | 3 | 3.9% | |
| - 45-54 | 7 | 5.6% | 3 | 3.9% | |
| - 55-64 | 3 | 2.4% | 2 | 2.6% | |
| - 65-74 | 3 | 2.4% | 1 | 1.3% | |
| - ≥75 | 4 | 3.2% | 2 | 2.6% | |
| - Total | 124 | 100.0% | 76 | 100.0% | |

[a]p-values calculated with chi-squared tests, excluding unknown values. Based on inclusion of unknown values, p=<0.001 for both age and sex.

Supplementary Figure 1. Estimates of the proportion of total influenza cases exposed to international travel, based on the subset of interviewed cases, Victoria, Australia, 1 November 2021 – 6 April 2022.

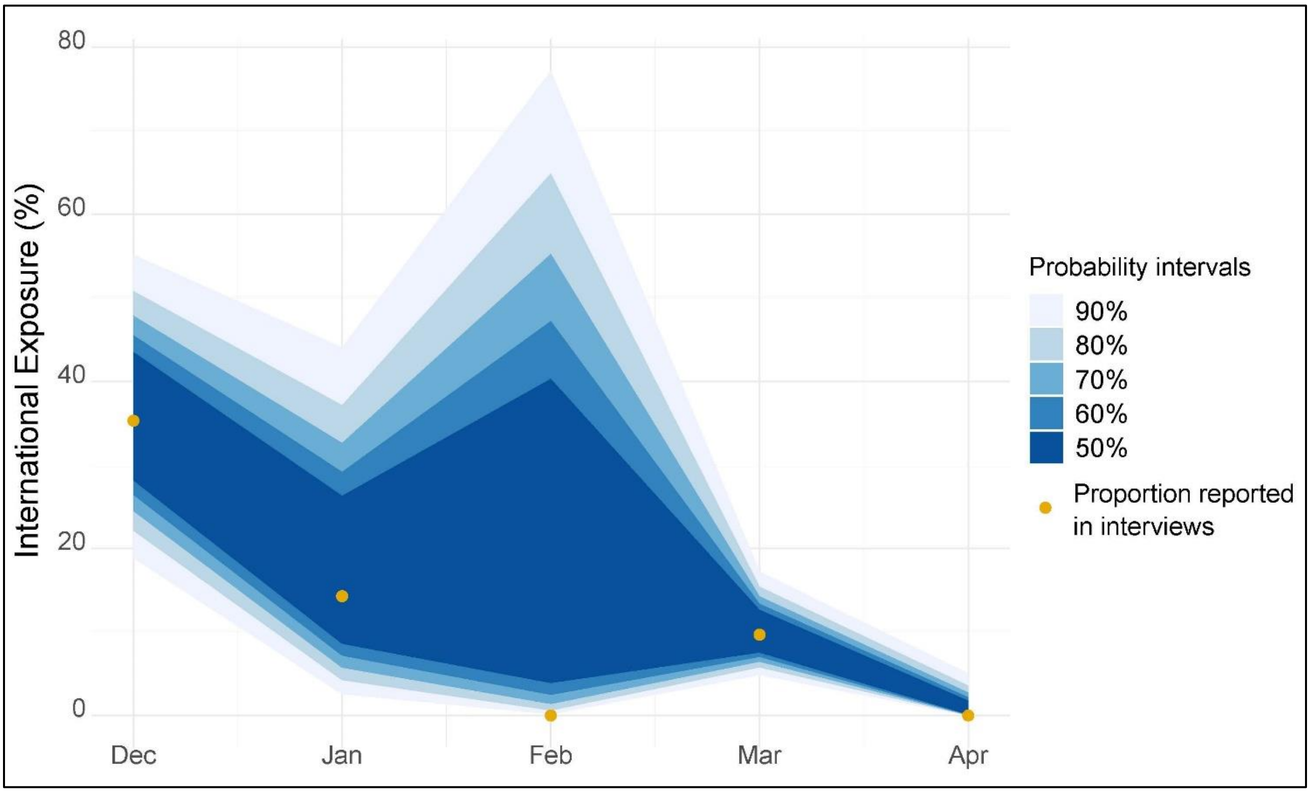


The plot shows estimates of the proportion of total influenza cases exposed to international travel, based on the subset of interviewed cases across different probability intervals. Analysis based on assumption that proportion of interviewed cases reporting international travel exposure is a binomial process. Data source: Department of Health and interviews conducted in this study.

Supplementary table 6. Health service attendance and pathways to accessing influenza testing data from case interviews for first 200 influenza notifications (n=124), Victoria, Australia, 1 November 2021 – 6 April 2022

| | First accessed | | Tested | | Accessed | |
|---|---|---|---|---|---|---|
| | n | % | n | % | n | % |
| GP | 55 | 45.1 | 43 | 35.2 | 66 | 54.1 |
| ED | 30 | 24.6 | 52 | 42.6 | 56 | 45.9 |
| COVID testing centre | 23 | 20.5 | 16 | 13.1 | 26 | 21.3 |
| Ambulance | 2 | 1.6 | 0 | 0.0 | 9 | 7.4 |
| Nurse-on-call[a] | 5 | 4.1 | 0 | 0.0 | 7 | 5.7 |
| Workplace | 2 | 1.6 | 3 | 2.5 | 3 | 2.5 |
| Specialist outpatient | 1 | 0.8 | 1 | 0.8 | 1 | 0.8 |
| Inpatient | 1 | 0.8 | 4 | 3.3 | 3 | 2.5 |

[a]Nurse-led telehealth advice service.

Supplementary figure 2. Cluster diagram of influenza cases.

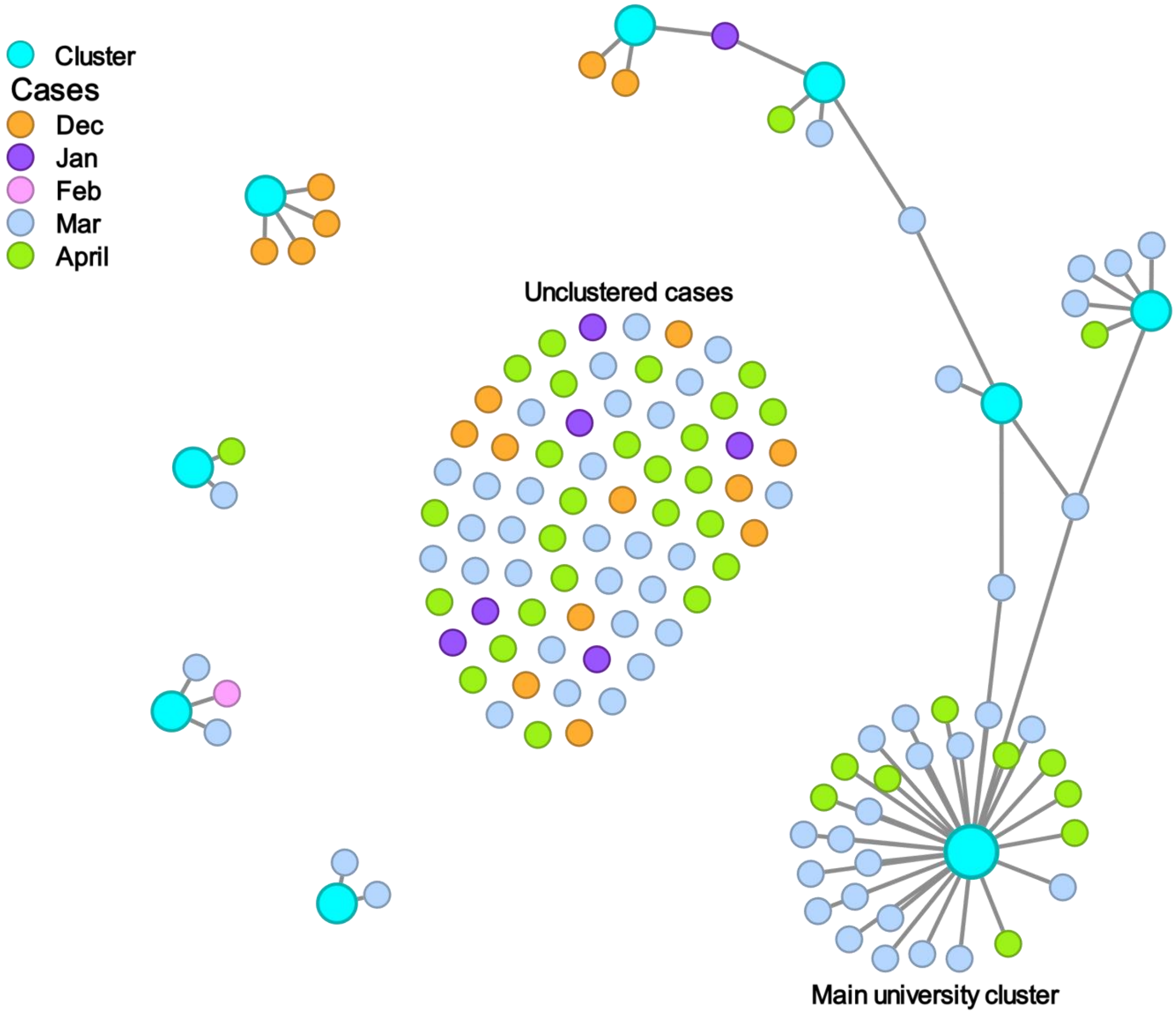


This figure is a representation of patterns of spread within the population. Clusters defined by common influenza exposures between interviewed cases. Only interviewed cases are depicted. Detailed contact tracing was not performed and in most instances it was not possible to identify direct person-to-person transmission between interviewed cases, based on these limitations. Data source: interviews conducted in this study.
